# The Role of Social Determinants of Health and Social Position in Mental Health: An Examination of the Moderating Effects of Race, Ethnicity, and Gender on Depression through the All of Us Dataset

**DOI:** 10.1101/2023.11.16.23298593

**Authors:** Matt Kammer-Kerwick, Kyle Cox, Ishani Purohit, S. Craig Watkins

**Affiliations:** The University of Texas at Austin

## Abstract

**Background:** Extant research has examined the roles of social position (SP) and social determinants of health (SDoH) on mental health outcomes. We add to this literature by focusing on major depressive disorder, investigating how race, ethnicity, gender, and sexual identity moderate the role of several social determinant domains on this common mental health condition.

**Methods:** Our analysis is based on the All of Us (AoU) dataset. We use a staged multiple logistic regression design. In the first stage, we consider how SP factors independently predict risk for diagnosis of MDD. In the second stage, we consider how SDoH add information to predict diagnosis of MDD. In the third stage, we consider how select SP factors moderate the role of SDoH in assessing risk for MDD diagnosis. We choose to focus on race/ethnicity and gender/sexual identity as SP moderators. We examine those moderating effects on food insecurity, discrimination, neighborhood social cohesion, and loneliness.

**Results:** Our findings further illustrate the complexity and nuance associated with how the context of where and how people live their lives has significant differential impact on health outcomes. Some of our results confirm long-standing relationships while elucidating detail about the effect on health. For example, independent of discrimination, Black community members have the same likelihood of an MDD diagnosis as Whites (AOR = 1.00, p = 0.982). However, discrimination experienced by Black community members increases their likelihood of a diagnosis of MDD (AOR = 1.47, p = 0.053) whereas among Whites experiencing discrimination does not increase the likelihood of an MDD diagnosis (AOR = 1.25, p = 0.122). Our analysis indicates that increases to loneliness for cisgender heterosexual female community members and gender and sexually minoritized community members are associated with lesser increases in risk of MDD diagnosis than similar increases in loneliness for cisgender heterosexual males (AOR = 0.44 and 0.22, p < 0.001, respectively), suggesting that this specific SDoH may have differential impacts across population segments. Other results shed new light on less well-established moderation effects. For example, gender and sexually minoritized community members are much more likely to experience depression compared to cisgender heterosexual men (AOR = 2.66, p < 0.001). Increasing neighborhood social cohesion does not alter the likelihood of depression, holding all other factors constant (AOR = 0.84, p = 0.181). But there is a weak moderation effect (AOR = 1.41, p = 0.090).

**Conclusions:** We use these analyses to outline future research to delve deeper into these findings. The current study demonstrates the value of the AoU data in the study of how various SDoH factors differentially drive health outcomes. It also provides a reminder that even larger datasets designed to represent the general population face substantial challenges for research focused on marginalized community segments and is a timely reminder that sampling plans are needed to ensure sufficient statistical power to examine those most marginalized and underserved.

## Introduction

In 2008, the WHO codified notions of health equity and “the conditions in which people are born, grow, live, work, and age” as the social determinants of health (SDoH) [1]. In the intervening nearly 15 years significant scholarship and policy efforts have been devoted around the globe to improve understanding and best practices for achieving the envisioned improvement in health equity based on therapeutic area, patient population, and various typologies for the determinant domains included. The WHO currently lists the following as “examples of the social determinants of health, which can influence health equity in positive and negative ways: income and social protection; education; unemployment and job insecurity; working life conditions; food insecurity; housing, basic amenities, and the environment; early childhood development; social inclusion and non-discrimination; structural conflict; and access to affordable health services of decent quality [2]. U.S. Department of Health and Human Services’ Healthy People 2030 program [3] states that SDoH “can be grouped in to 5 domains: economic stability, education access and quality, healthcare access and quality, neighborhood and built environment, and social and community context.” These two perspectives serve to illustrate that there are different taxonomies for SDoH. For the interested reader, in their study of diabetes, Hill-Briggs and colleagues (2021) compare and contrast the SDoH schema by the WHO, Healthy People 2020, County Health Rankings Model (2014), and Kaiser Family Foundation (2018) [4].

Recent research reveals the significant role of SDoH factors in moderating relative risk of developing chronic diseases such as cardiovascular disease, diabetes, cancer, and mental health [4–6]. Researchers, for example, have examined low socioeconomic status and unsafe neighborhoods are risk factors that make some populations more susceptible to developing chronic illnesses. Understanding and addressing social determinants of health is crucial in promoting health equity and reducing health disparities in marginalized populations in order to ensure all individuals have the opportunity to live healthy, fulfilling lives.

The current study probes the effects of SDoH in relation to major depressive disorder (MDD). MDD is one of the most common forms of mental health disorders in the U.S. The National Institute of Mental Health defines MDD as, “a period of at least two weeks when a person experienced a depressed mood or loss of interest or pleasure in daily activities, and had a majority of specified symptoms, such as problems with sleep, eating, energy, concentration, or self-worth.” According to the NIMH, 21 million U.S. adults, or 8.4% of all adults, had at least one major depressive episode in 2020 [7]. The distribution of MDD varies noticeably among adults in the U.S. For example, from the same report, women (10.5%) have higher levels of MDD compared to men (6.2%). Similarly, young adults ages 18-25 suffer higher rates of MDD than their older counterparts aged 50 and older (5.4%). There are also racial and ethnic differences in the prevalence of MDD among U.S. adults. In terms of race and ethnicity, persons who report two or more racial/ethnic identities have the highest (15.9%) prevalence of MDD compared to whites (9.5%), Latinx (7.0%), Blacks (6.0%), and Asians (4.2%) [7].

## Background

To set the stage for the current study, we review depression, as exacerbated by COVID and as an enduring societal and health challenge; the complex connections between various SDoH and depression; and the resources available in the All of Us dataset [8] that we utilize to probe SDoH in relation to depression.

### Depression

The COVID-19 pandemic has greatly impacted the mental health of millions of people around the world, leading to accelerated rates of MDD. In the U.S. rates of MDD have risen sharply as a result of COVID-induced societal changes including, for instance, greater social isolation and loneliness [9] and economic stress due, in part, to changes in the nature and availability of work [10]. Concerns about the long-term health impacts of the coronavirus seem tied to experienced COVID severity with gender playing a moderating role [11,12]. The pandemic also exposed the impacts of systemic inequality in health outcomes [13]. The National Center for Health Statistics (NCHS) partners with the U.S. Census Bureau to produce the Household Pulse Survey, a reoccurring survey instrument that measures, among other things, the percentage of adults who report symptoms of anxiety or depression that have been shown to be associated with diagnoses of generalized anxiety disorder or major depressive disorder. This online survey was designed to support the ability of the federal government to capture rapid and longitudinal data in order to provide relevant information about the impact of the coronavirus pandemic [7].

In December of 2022, the Household Pulse Survey revealed that 23.6% of those surveyed by the NCHS reported MDD symptoms [7]. Additionally, the same study suggests that while the mental health of many U.S. adults suffered during the pandemic, the rate of decline among marginalized populations was especially acute. Nearly a quarter of women (24.9%) reported MDD symptoms compared to 22.1% of men. And whereas 21.2% of persons who identified as “straight” reported MDD symptoms the rate of self-report among persons who identify as “bisexual” (43.4%) or “gay and lesbian” (33.5%) is notably higher. The NCHS survey also found sharp racial differences in the self-reporting of MDD during the coronavirus pandemic. Persons who identified as Asian (17.9%) had the lowest self-reports among all racial and ethnic groups selected for the survey compared to Latinx (26.4%), Blacks (25.8%), and Whites (22.3%). Persons who identified with more than one race had the highest self-reported rates of MDD (33.4%).

Beyond the role of COVID, these data suggests that MDD is connected to broader social, cultural, and population factors. In this paper we focus on SDoH to identify the unique social-structural factors that are associated with an MDD diagnosis. Moreover, the current study investigates how race, ethnicity, and gender moderate the role of several social determinant domains on depression. Our analysis is based on the All of Us dataset, a National Institutes of Health (NIH) asset designed to facilitate analysis of a broad range of health conditions and types of data including electronic health records, genomic, wearable, and, for our purposes, SDoH. All of Us also spans the breadth, depth, and diversity of the United States population [8].

The varied dimensions of mental illness—symptoms, cognitive impairments, and recurrences—can be severe for the individuals and families who experience a condition. MDD, for example, is linked to mortality. The mortality risk increases by 60-80% for people with MDD [14]. The risk may be attributed to suicide, but recent studies have shown MDD to increase the risk for cardiovascular death and stroke as well [15]. Moreover, there are significant economic impacts associated with depression. Depression is associated with lowered income or unemployment attributed to decreased workplace productivity and increased absenteeism [16]. Workplace-related absenteeism attributed to MDD in the US has been estimated to result in a loss of $36.6 billion every year to the economy [17].

### SDoH and Depression

While there is widespread consensus that SDoH influence health outcomes we need more empirical-based studies to explicate the nuances of these interactions. Not all SDoH are necessarily equal in prevalence, distribution, or impact. For example, it is highly likely that the distribution and impact of SDoH will vary significantly by population characteristics. Women with children, for example, may be more vulnerable to certain SDoH than women without children. In other words, we need to begin understanding with more precision how, when, and for whom specific SDoH matter. This paper addresses some of these issues by examining the moderating effects of population characteristics like race/ethnicity, gender, and age on the association between SDoH and major depression disorder (MDD). As Alegria et al. (2018) note, scholars need to investigate the differential effects of SDoH on members of different populations [18]. In that study, Alegría and colleagues summarized a list of SDoH domains that have been found to be involved with deleterious mental health outcomes, including minorized race/ethnicity, immigration/migration status, sexual orientation, gender identity, and nationality.

Oher studies have also found an association between SDoH and mental illness. Gnanapragasam et al (2021) note the important social determinants of mental health to include, among other things, adverse education, employment, poverty, food insecurity, housing and built environment and discrimination [19]. Similarly, according to Compton and colleagues (2015), the core social determinants of mental health include “racial discrimination and social exclusion; adverse early life experiences; poor education; unemployment, underemployment, and job insecurity; poverty, income inequality, and neighborhood deprivation; poor access to sufficient healthy food; poor housing quality and housing instability; adverse features of the built environment; and poor access to health care” [20]. Jeste and Pender (2022) additionally note, “homelessness, social isolation with associated loneliness, social media, positive childhood experiences, social connections, and community-level resilience” as factors that exacerbate outcomes for patients with existing serious mental illnesses and substance use disorders [21]. Adverse social conditions trigger stress responses and extended exposure to them may detrimentally affect mental health outcomes [22].

Discrimination, food insecurity, and neighborhood social cohesion may be important causes of physical and mental health disparities among populations with intersectional social identities. For instance, a study on food insecurity, depression, and race examined the prevalence of food insecurity among 131 students aged 18 to 24 at a college in Mississippi [23]. The US Department of Agriculture’s Household Food Security Survey Module: Six-Item Short Form was used to test food security, and the Patient Health Questionnaire-9 was used to measure depression. According to the findings, African American pupils were more likely to experience food insecurity than Caucasian students. Students who experienced severe food insecurity had a 4.52-times higher likelihood of developing depression than those who did not [23].

Moreover, the relationship between neighborhood cohesion and psychological distress among racial/ethnic-sexual orientation groups was also examined in a study using zero-order multinomial logistic regression models. Drawing on a body of extant literature, the authors define neighborhood cohesion as a feeling of connectedness to one’s neighborhood that facilitates the development of supportive relationships [24]. Results showed neighborhood cohesion provided greater protection for heterosexual groups against moderate psychological distress compared to their non-heterosexual counterparts. A difference of 8.16 predicted probability was found for moderate psychological distress for white non-LGB individuals compared to a difference of 5.14 for LGB white individuals. Further, a predicted probability of 4.87 for moderate distress was found for heterosexual Black individuals when compared to a 3.39 increase for LGB Black and a difference of 5.14 predicted probability for Latinx versus 2.14 for LGB Latinx [24]. Further, neighborhood cohesions also seemed to be a significant protective factor against severe psychological distress for lesbian, gay, and bisexual groups [24]. This can be seen in predicted probabilities for severe psychological distress for LGB white was 5.26 while it was 3.41 for heterosexual white. Further, predicted probability for severe psychological distress for LGB Black was 1.69 as compared to 1.59 for non-LGB Black and 4.66 for LGB Latinx compared to 2.61 non-LGB Latinx [24]. One possible reason for this may be the idea that lesbian, gay, and bisexual white individuals experience the greatest protective effect from neighborhood cohesion from the combined effects of protective influences in more cohesive neighborhoods.

Swann et al. (2020) found that enacted stigma based on race/ethnicity and sexual/gender minority (SGM) status was a significant predictor of mental health outcomes as well as alcohol-related problems. They evaluated the effects of "racial discrimination, SGM victimization, and sexual orientation microaggressions on depression symptoms, anxiety symptoms, alcohol-related problems, and marijuana-related problems" [25]. In the sample that was examined, it was discovered that 45.1% of participants said they had not experienced victimization in the six months prior, as opposed to 2.3% who said the same about microaggressions [25]. This may be explained by the assumption that victimization is less frequent than more subtly harmful stigmatizing behaviors like microaggressions, which may affect a larger number of SGM and be more detrimental to mental health.

Further, a study evaluating multifactorial discrimination as a fundamental cause of mental health disparities used baseline data from the Project STRIDE: Stress, Identity, and Mental Health to find multifactorial discrimination was a significant risk factor for high depression scores in terms of chronic strain and total number of stressful life events as noted by the adjusted odds ratio of 32.30 and 1.04 respectively [26]. Multifactorial discrimination may be defined as “the total discrimination experienced by such individuals, accounting for their sexual orientation, gender, race/ethnicity, and/or other identity markers” [26]. Results found women to be more likely to have high depression along with Latino/Hispanics groups suggesting patterns of poor health differed by sex and race/ethnicity. Further, subjects with low depression scores showed anxiety and aggregate mental health scores that were influenced by discrimination [26].

### Examination of Social Position, SDoH, and Depression through the All of Us Dataset

The complex interplay between objective sociodemographic factors and subjective assessments of social position (SP) has been studied without universal agreement on definitions; see Alder and colleagues (2000) for a discussion [27]. Lindemann (2007) examined an individual’s perceived place in society, or social position, and the influence of observable variables like age, gender, ethnicity, education, employment, and income. As Lindemann states, “subjective social position depends not only on the objective characteristics but also on how people experience society, the way they perceive their position in comparison with others, and what they imagine their position would be in future.” [28] We follow the lead of Avlund et al (2003), who included education, occupation, social class, income, and housing tenure as measures of social position in a study of depression, finding strong associations between social position and measures of health [29]. In this paper we use the term social position in relation to such sociodemographic labels to underscore the fact that they have social aspects and societal implications for the persons who experience them. The All of Us Research Program includes a survey module called The Basics that includes these variables.

The All of Us Research Program explicitly addresses the need for greater diversity in medical research [8,30]. Mapes et al (2020) review the scope of the diversity design in the program and describe efforts made to ensure that participants reflect the diversity of the US [31]. The All of Us Research Program added its SDoH questionnaire in its third year [32]. As such, its design was additive to the surveys implemented earlier that include questions germane to SDoH. Additionally, the sample base of participants who have completed the SDoH survey is a subset of the total sample. The All of Us SDoH survey consists of 81 questions which are drawn from several different instruments that are listed in the Supplement. Additional details about these instruments are available at the All of Us website.

Barr and colleagues assessed the prevalence of various psychiatric diagnoses in the All of Us dataset compared to other published estimates [33]. There are multiple numbers of mental health diagnosis in the All of Us dataset including, for example, MDD, bipolar, generalized anxiety disorder, and PTSD. Their comparison utilizes phecodes available in All of US to label diagnoses. As explained in their measures section, diagnoses for disorders were based on phecodes derived from billing codes of the International Statistical Classification of Diseases, Ninth and Tenth Revisions, Clinical Modification (ICD-9/10-CM). Individuals with 2 or more phecodes from the selected CD-9/10-CM codes under the broader categories of drug-related disorders, mental disorders, substance abuse, sleep disorders, and mental state, were considered to have a diagnosis.

Further, Barr and colleagues [33] examine disparities across sociodemographic factors that were ultimately found to be similar to disparities from nationally representative samples. Non-Hispanic White, female at birth, women, and LGBTQ individuals in the sample were at increased risk of most disorders. However, Black, multiracial, and other non-White participants were particularly susceptible to schizophrenia which may be attributed to racism and biases in diagnoses. Having a college degree and an annual household income more than $100,000 served as protective factors that decreased risk for each disorder. Their paper concludes with positive endorsement of the robustness of the AoU data relative to the general population.

### The Current Study

These considerations led our team to address the following research questions in our analysis of the All of Us dataset:

> RQ1. To what degree, if at all, does social position predict risk of depression?
>
> RQ2. To what degree, if at all, do SDoH factors predict risk of depression?
>
> RQ3. How does social position moderate the role of social determinants of health on risk of depression?

Specifically, to address health equity, in RQ3, we focus on the moderating effects of race/ethnicity and gender identity/sexual orientation while controlling for other social position factors. Based on our review of the literature, we examine those moderating effects on food insecurity, discrimination, neighborhood social cohesion, and loneliness.

The paper proceeds as follows: we provide additional information about the All of Us design and how we operationalize the social position variables used in the models and the aggregation scheme employed in our analyses; how we selected and operationalized the SDoH measures from the add-on SDoH survey plus select measures from the other surveys; and how we defined and operationalized our response variables for depression from the All of Us database of medical concepts from participant electronic health records. We then specify the models we employed to answer our research questions and present the results of our analysis using those models. The paper concludes with a discussion of our findings, limitations of the current study, and future research opportunities.

## Method

We developed a computational workflow in the AoU workbench environment to query tables to create an analytical dataset. In this section we document how we operationalized features for our analysis. Our methods are documented and available from the lead author on the AoU platform.

### Response Feature Operationalization

Our study considers the outcome Major Depressive Disorder (MDD). 392,260 participants contributed electronic health records to All of Us, which populate the medical concepts database. We use the medical concepts “Major depressive disorder” (SNOMED code 370143000) as listed in the All of Us data browser.

Of those contributing electronic health records with at least one condition recorded in the database, 26.2% recorded the MDD SNOMED code in their records. The MDD code refers to a hierarchy of conditions that includes single episodes of major depression, recurrent major depression, major depression in remission, minimal major depression, mild major depression, moderate major depression, severe major depression, major depression with psychotic features, and melancholic-type major depression. Our analytical framework organizes the data matrix into individual records corresponding to participants and variables representing each feature or outcome label. Within this framework, MDD status is binary-encoded.

### Social Position Feature Operationalization

All social position features were sourced from The Basics survey, which all participants are required to complete. Five factors were selected for our analysis: gender/sexual identity, household income, education level, race/ethnicity, and home ownership. These categories are further decomposed into the following features:

1. Gender/sexuality: cisgender heterosexual male, cisgender heterosexual female, LGBTQIA2+ This feature category is a composite of the gender and sexual orientation questions in The Basics survey. Participants were allowed to choose multiple options for the sexual orientation question. LGBTQIA2+ includes those who identified as one or more of the following: bisexual sexual orientation, gay sexual orientation, lesbian sexual orientation, no sexual orientation, nonbinary gender, transgender, or additional options for gender. Cisgender heterosexual male includes only those who identified as straight and male, and cisgender heterosexual female includes only those who identified as straight and female.
2. Income: greater than or equal to $35,000 per year, less than $35,000 per year Answers for the household income question were binned into 9 groups: less than $10,000; $10,000-$24,999; $25,000-$34,999; $35,000-49,999; $50,000-$74,999; $75,000-$99,999; $100,000-$149,999; $150,000-$199,999; $200,000 or more. We binary encode this feature by choosing a $35,000 threshold.
3. Education: college degree, no college degree Answers for the highest grade of education completed are binned into 8 groups: never attended, grades 1-4, grades 5-8, grades 9-11, grade 12 or GED, one to three years of college, college graduate, advanced degree. We binary encode this feature into college graduate and not a college graduate labels. 66.8% of those in our analytical sample reported college graduate status.
4. Race/ethnicity: White, Hispanic/Latinx, Black or African American, Asian, more than one population, or other. In The Basics survey, All of Us participants were asked to select which racial-ethnic categories describe them. Participants were allowed to select multiple answers to this question. Participants who selected more than one answer are categorized as more than one population. In our analytical sample, Middle Eastern/North African and Native Hawaiian/other Pacific Islander responses are combined to form the “Other” race/ethnicity category.
5. Home ownership: homeowner, not a homeowner The All of Us survey question regarding living arrangement offers three response choices: ownership, rental, or other arrangement. We aggregate the rent and other arrangement labels to create just two labels: homeowner and not a homeowner.
6. Age: older than 65, younger than 65 There is no age field natively encoded in All of Us. However, there are fields for date of birth and time at which a survey was taken. For our analytical sample, we consider the age at which a participant took the Social Determinants of Health survey, in years. Individuals younger than 18 years are not permitted to participate in All of Us. The six social position labels are factorized into their respective categories. To prevent collinearity, each label is designated a reference category, which is omitted from the social position feature set for regression. After removing the reference features from the feature set, we are left with eleven social position features. For all of the above fields, if an individual recorded a skipped question, preferred not to answer, or otherwise did not answer a question, they are not included in the analytical sample. The distribution of social position data is presented in Supplement 2. The prevalence of MDD by social position is presented in Table 1.

**Table 1.**
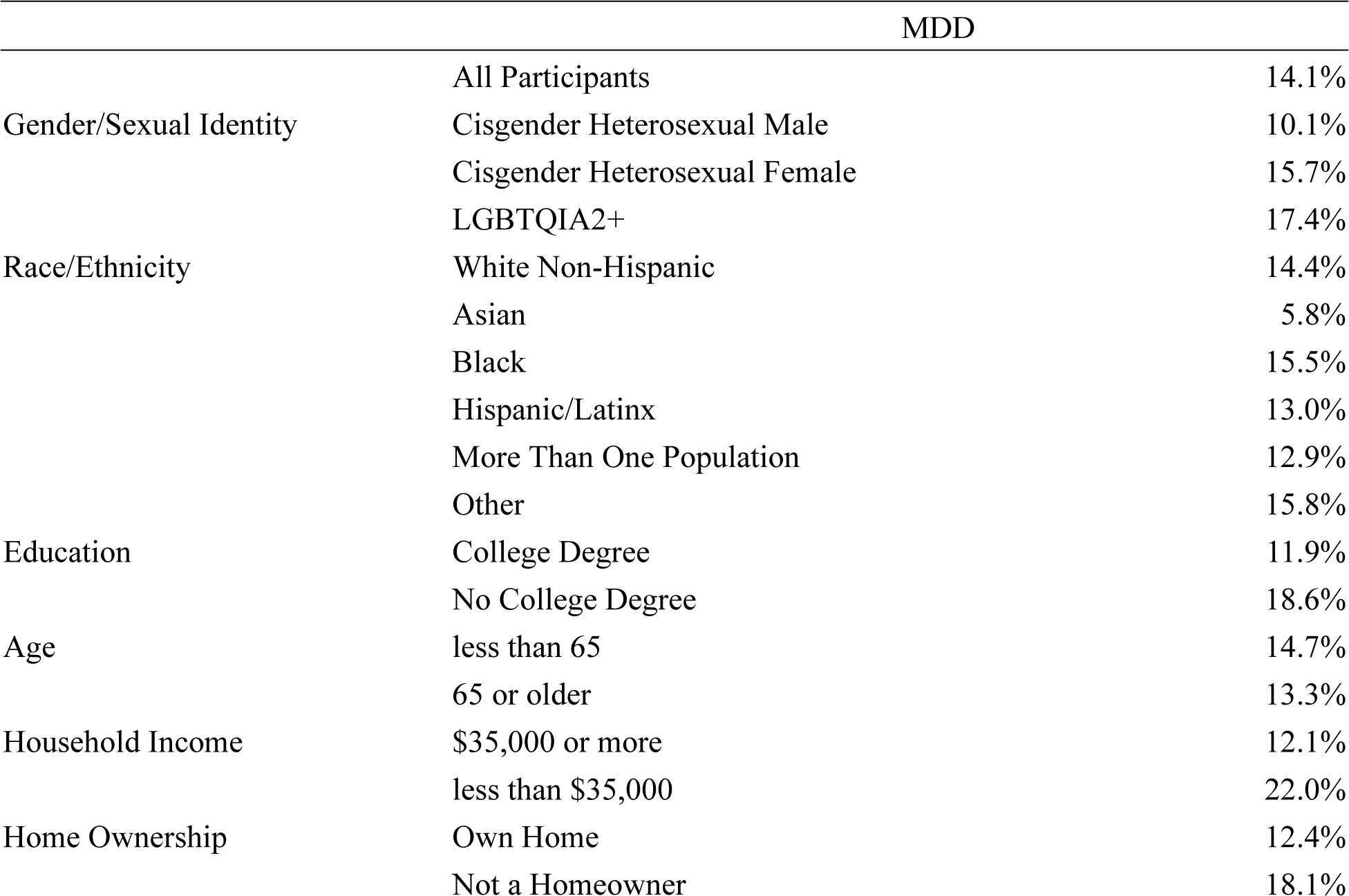
Analytical Sample MDD Prevalence.

### SDoH Feature Operationalization

All of Us Social Determinants of Health Survey questions were aggregated from several validated instruments designed to measure specific social determinants of health. However, health care access and coverage are not captured in the All of Us Social Determinants of Health survey. Instead, these items are covered in The Basics and Health Care Access and Utilization surveys, which were administered before the Social Determinants of Health Survey. Informed by a review of extant SDoH literature [18,34–37] and the AoU design, we combined selected health care questions from these other surveys with the Social Determinants of Health survey to form the corpus of questions for our analysis of social determinants of health. These questions are partitioned into ten categories.

We partitioned the Social Determinants of Health Survey questions by their source instrument and treated each source instrument as a unique social determinant of health field. However, some source instruments were excluded because of thematic overlap with other instruments. The “Discrimination in Medical Settings” survey was dropped due to its similarity to the “Everyday Discrimination” survey. Additionally, the “RAND MOS Social Support Survey Instrument” was dropped due to its to the UCLA Loneliness Scale. The “Cohen’s Perceived Stress Scale” was also dropped because of its similarity to anxiety screening tools and the comorbidity of anxiety disorders and MDD. The English proficiency questions from the California Health Interview Survey were also dropped because there were few non-fluent English speakers in our analytical sample. Lastly, the “Brief Multidimensional Measure of Religiousness/Spirituality” survey was dropped because religion is rarely included as a social determinant of health.

In summary, we use ten social determinants of health features.

1. Food Security
2. Discrimination
3. Neighborhood Disorder
4. Neighborhood Social Cohesion
5. Neighborhood Infrastructure/Facilities
6. Number of Moves in the Past 12 Months
7. Loneliness
8. Housing Issues
9. Lack of Health Care Access
10. Health Insurance

There are four different encoding types for the social determinants of health fields: mean subscales, numeric responses, sum of checked responses, and indicator; see Table 2. These are further described below.

**Table 2.**
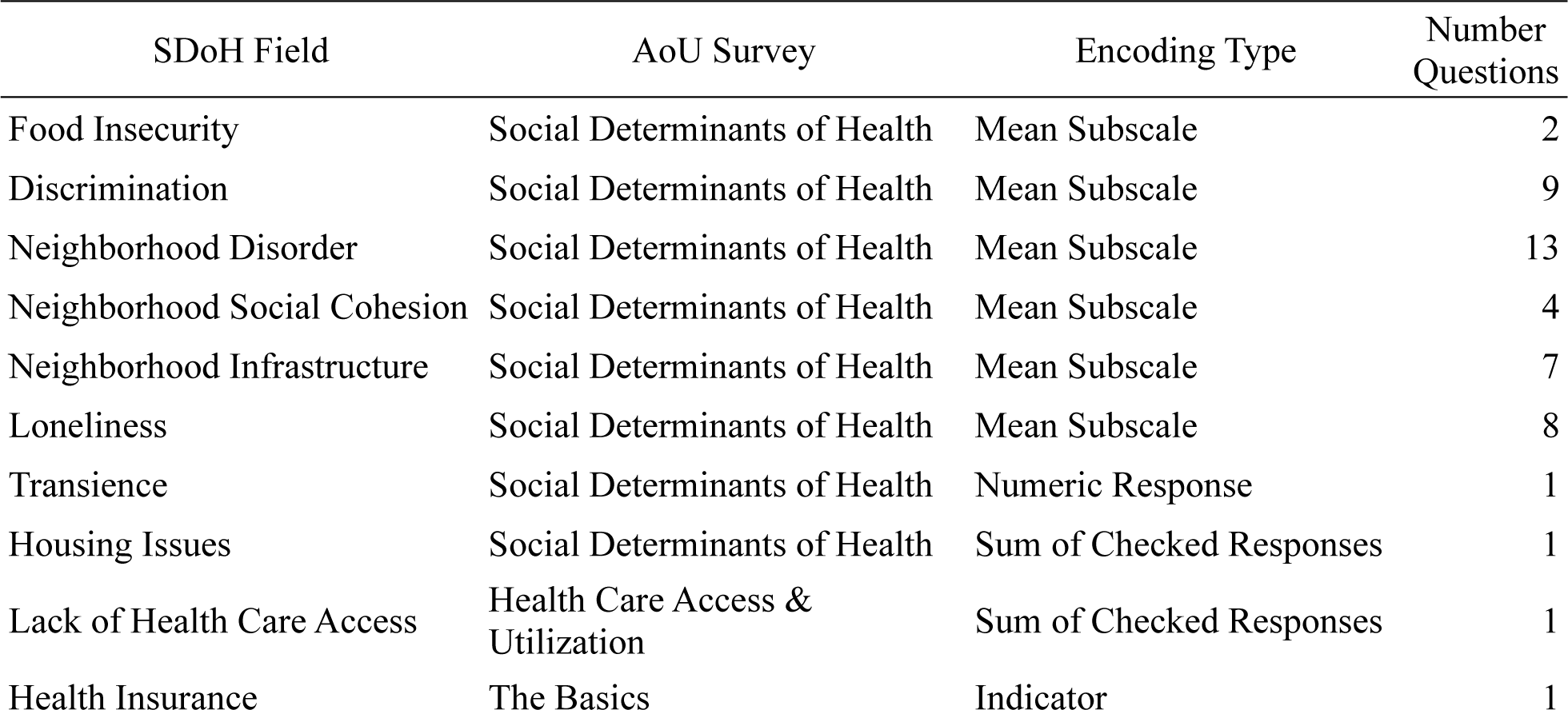
Summary of SDoH Factors with Source, Encoding, and Number of Questions.

### Data Type 1: Mean Subscale

The mean subscale encoding reflects the fact that our social determinants of health fields partition survey questions by the unique survey instruments from which they were sourced. Rather than use the response to each individual question in our analysis, we create a single score for each sub-survey, allowing us to represent each social determinant of health field with a single value. This is performed as follows:

1. Likert scale responses are encoded as integer values 1 through *n*, where *n* is the number of possible responses on the scale. For example, for the question asking, “How much you agree or disagree that your neighborhood is clean?” the responses strongly disagree, disagree, agree, and strongly agree are mapped to 1, 2, 3, and 4 respectively.
2. Instances where participants skipped, preferred not to answer, or otherwise did not answer a question are recorded as null values in the dataset.
3. Questions within each field originate from a consistent source and were thematically aligned. Generally, the questions in a given field share a similar valence where higher scale values represent positive outcomes and lower values denote negative outcomes. In instances where answer scales diverge in valence, the values for certain questions are inverted to ensure uniformity in the direction of encoding across all questions within the field.
4. All scaled responses are re-scaled using a min-max standard scaler so that their values fall between 0 and 1.
5. Each field contains some number of questions, but some participants did not respond to all the questions in a given field. If a field contains three or fewer questions, we require participants to respond to all questions in the field to be included in the analytical sample. If a field contains more than three questions, we require participants to respond to at least three questions to be included in the analytical sample. We set this threshold to allow participants who did not answer all of the questions in a given field to remain in the analytical sample.
6. We let each field equal the mean of the responses to its questions. Because question responses are re-scaled to fall between 0 and 1, each field is equal to a value between 0 and 1. Null values are omitted from the mean value calculation.

### Data Type 2: Numeric Response

The Transience field is encoded as a numeric response from a single question: “In the last 12 months how many times have you or your family moved from one home to another?” We simply take the response to this question as the field value.

### Data Type 3: Sum of Checked Responses

Two fields are encoded as sums of checked responses: Housing Issues and Lack of Health Care Access. These fields reflect the response to one question each. These questions accept “check all that apply” responses. We take the sum of the number of checked responses as the field value. For example, the Housing Issues field uses the question from the Social Determinants of Health Survey, “Think about the place you live. Do you have problems with any of the following?” Excluding the “None of the above response,” we sum the number of checked responses, indicating the number of relevant problems in the home, and take that as the field value. These values are then re-scaled by the min-max scaler.

### Data Type 4: Indicator

Only the Health Insurance field takes an indicator value. In this field, 1 represents an affirmative response to the question, “Are you covered by health insurance or some other kind of health care plan?” and 0 represents otherwise.

After removing those participants who did not take the Social Determinants of Health survey or had null values in their social position or SDoH fields, we are left with an analytical sample of size 86,960.

## Procedure and Analysis Strategy

We use a staged multiple logistic regression design. In the first stage, we consider how social position factors independently predict risk for diagnosis of MDD. In this stage, the feature set consists of only the eleven social position features. In the second stage, we consider how social determinants of health add information toward prediction of MDD diagnosis. In this stage, the feature set consists of the 11 social position features and the 10 social determinants of health features. In the third stage, we consider how select social position factors moderate the role of social determinants of health in predicting MDD diagnosis. In this stage, the base feature set consists of the union of social position and social determinants of health features, of size 21. We choose to focus on race/ethnicity and gender/sexual identity as social position moderators. We also exclude the age, health care access, and health insurance social determinants of health from consideration as moderation effects, instead interpreting these as controls, as access to health care and availability of health insurance are bottlenecks to receiving a diagnosis of MDD. We then consider pairwise interaction terms between selected social position moderators and social determinants of health. In RQ3, we choose to focus on the moderating effects of gender/sexual identity and race/ethnicity on food insecurity, discrimination, neighborhood social cohesion, and loneliness. The set of interaction terms examined in RQ3 is of size 28. We further use expected marginal means interaction analysis to isolate specific interaction effects and corroborate findings from the regression models. Table 3 summarizes our modeling approach and the features used in each stage of our analysis.

**Table 3.** Feature Sets.

|  |
| --- |
| Dependent Variables |
| Indication (0/1) for each of: |
| Major Depressive Disorder (MDD) |
| Stage 1: Social Position – Research Question 1 |
| Age (Reference = Age $\geq 65$)<br>Gender/Sexual Identity (Reference = Cisgender Heterosexual Male, 3 levels)<br>Minority Race/Ethnicity (Reference = White Non-Hispanic, 6 levels)<br>Education (Reference = College Degree or More, 2 levels)<br>Poverty (Reference = HHI $\geq \$35,000$, 2 levels) |
| Stage 2: Social Determinants of Health - Research Question 2 |
| Food Security (Calculated factor scaled between 0 – 1)<br>Discrimination (Calculated factor scaled between 0 – 1)<br>Neighborhood Disorder (Calculated factor scaled between 0 – 1)<br>Neighborhood Social Cohesion (Calculated factor scaled between 0 – 1)<br>Neighborhood Infrastructure/Facilities (Calculated factor scaled between 0 – 1)<br>Number of Moves in Past 12 Months (Transience) (Calculated factor scaled between 0 – 1)<br>Loneliness (Calculated factor scaled between 0 – 1)<br>Housing Issues (Calculated factor scaled between 0 – 1)<br>Health Care Access (Calculated factor scaled between 0 – 1)<br>Health Insurance (Calculated factor scaled between 0 – 1) |
| Stage 3: Select Social Position Moderators - Research Question 3 |
| Food Insecurity X:<br>Gender/Sexual Identity (Reference = Cisgender Heterosexual Male)<br>Minority Race/Ethnicity (reference = White Non-Hispanic)<br>Discrimination X:<br>Gender/Sexual Identity (Reference = Cisgender Heterosexual Male)<br>Minority Race/Ethnicity (reference = White Non-Hispanic)<br>Neighborhood Social Cohesion X:<br>Gender/Sexual Identity (Reference = Cisgender Heterosexual Male)<br>Minority Race/Ethnicity (reference = White Non-Hispanic)<br>Loneliness X:<br>Gender/Sexual Identity (Reference = Cisgender Heterosexual Male)<br>Minority Race/Ethnicity (reference = White Non-Hispanic) |

## Results

Table 4 reports an overview of the results of hierarchal logistic regression model. The table includes fit statistics for each stage of the model as information is added to the hierarchy. The Akaike information criteria (AIC) for each model, the change in the *X*^2^ statistic as information is added, and the significance of the change in model fit are presented.

**Table 4.** Hierarchical Modeling Summary.

| Model Stage | df | AIC | $\Delta X^2$ | $\Delta X^2$ df | Sig. |
| --- | --- | --- | --- | --- | --- |
| Intercept | 1 | 70842.63 |  |  |  |
| Social Position | 12 | 68768.45 | 2096.18 | 11 | <0.001 |
| + SDoH | 22 | 67387.91 | 1400.54 | 10 | <0.001 |
| + Moderation | 50 | 67309.24 | 134.6657 | 28 | <0.001 |

Table 5 reports the results for Stage 1, social position; Stage 2 (+ SDoH), and Stage 3 (+ select moderations for RQ3). Few parameter estimates change substantively as new information is added to the model. For example, between Stages 1 and 2, only Age Older than 65 changed significantly, with a Stage 1 AOR estimate of 1.02, p = 0.363, that increased to 1.10, p < 0.001. For the sake of parsimony, we focus our discussion on parameter estimates that are significant at 0.01 in the Stage 3 model as well as estimates predicted to be significant in the literature but for which our results failed to find similar results.

**Table 5.** Detailed Model Results. Caption. Table 5 shows the main effects to address RQ1 and RQ2 as well as the interactions terms included in our analysis to assess the moderation effects associated with RQ3. Abbreviations: Cis Het = Cisgender Heterosexual, LGBTQIA2 = Lesbian, Gay, Bisexual, Transgender, Queer, Intersex, Asexual, Two Spirit, and additional minoritized gender and sexual identities.

|  | Stage 1 (Social Position) |  |  | Stage 2 (+ SDoH) |  |  | Stage 3 (+ Select Moderation) |  |  |
| --- | --- | --- | --- | --- | --- | --- | --- | --- | --- |
|  | AOR | CI | Sig. | AOR | CI | Sig. | AOR | CI | Sig. |
| **Social Position** |  |  |  |  |  |  |  |  |  |
| Constant | 0.09 | (0.09, 0.09) | <0.001 | 0.02 | (0.02, 0.03) | <0.001 | 0.02 | (0.01,0.02) | < 0.001 |
| Cisgender Heterosexual Female | 1.60 | (1.52, 1.67) | <0.001 | 1.52 | (1.45, 1.59) | <0.001 | 2.10 | (1.59,2.76) | < 0.001 |
| LGBTQIA2+ | 1.64 | (1.53, 1.76) | <0.001 | 1.43 | (1.33, 1.53) | <0.001 | 2.66 | (1.8,3.92) | < 0.001 |
| Income ≤ \$35,000 | 1.65 | (1.57, 1.74) | <0.001 | 1.42 | (1.35, 1.50) | <0.001 | 1.42 | (1.35,1.5) | < 0.001 |
| No College Degree | 1.41 | (1.35, 1.47) | <0.001 | 1.36 | (1.31, 1.42) | <0.001 | 1.37 | (1.31,1.43) | < 0.001 |
| Race/Ethnicity: Asian | 0.37 | (0.31, 0.45) | <0.001 | 0.38 | (0.32, 0.45) | <0.001 | 0.10 | (0.03,0.3) | < 0.001 |
| Race/Ethnicity: Black | 0.74 | (0.69, 0.80) | <0.001 | 0.78 | (0.72, 0.85) | <0.001 | 1.00 | (0.69,1.45) | 0.982 |
| Race/Ethnicity: Hispanic | 0.64 | (0.58, 0.69) | <0.001 | 0.72 | (0.66, 0.79) | <0.001 | 0.76 | (0.49,1.17) | 0.202 |
| Race/Ethnicity: Multiple | 0.75 | (0.67, 0.83) | <0.001 | 0.73 | (0.65, 0.81) | <0.001 | 0.72 | (0.4,1.29) | 0.269 |
| Race/Ethnicity: Other | 0.98 | (0.83, 1.16) | 0.812 | 0.92 | (0.78, 1.08) | 0.308 | 0.80 | (0.32,1.98) | 0.624 |
| Not a Homeowner | 1.26 | (1.20, 1.32) | <0.001 | 1.14 | (1.08, 1.19) | <0.001 | 1.13 | (1.08,1.19) | < 0.001 |
| Age Older than 65 | 1.02 | (0.98, 1.06) | 0.363 | 1.10 | (1.05, 1.15) | <0.001 | 1.11 | (1.06,1.16) | < 0.001 |
| **SDoH** |  |  |  |  |  |  |  |  |  |
| Food Insecurity |  |  |  | 1.25 | (1.13, 1.39) | <0.001 | 1.39 | (1.08,1.77) | 0.009 |
| Discrimination |  |  |  | 1.26 | (1.10, 1.45) | 0.001 | 1.25 | (0.94,1.67) | 0.122 |
| Neighborhood Disorder |  |  |  | 0.96 | (0.84, 1.11) | 0.586 | 0.95 | (0.83,1.1) | 0.515 |
| Neighborhood Social Cohesion |  |  |  | 0.92 | (0.81, 1.04) | 0.192 | 0.84 | (0.65,1.09) | 0.181 |
| Neighborhood Infrastructure |  |  |  | 1.04 | (0.95, 1.15) | 0.387 | 1.03 | (0.94,1.13) | 0.530 |
| Transience |  |  |  | 0.94 | (0.91, 0.96) | <0.001 | 0.93 | (0.91,0.96) | < 0.001 |
| Loneliness |  |  |  | 7.42 | (6.50, 8.46) | <0.001 | 15.09 | (11.48,19.83) | < 0.001 |
| Lack of Health Care Access |  |  |  | 1.31 | (1.17, 1.47) | <0.001 | 1.35 | (1.2,1.52) | < 0.001 |
| Health Insurance |  |  |  | 1.61 | (1.50, 1.74) | <0.001 | 1.64 | (1.42,1.89) | < 0.001 |
| Housing Issues |  |  |  | 0.97 | (0.89, 1.05) | 0.705 | 0.99 | (0.84,1.17) | 0.909 |
| **Select Interactions** |  |  |  |  |  |  |  |  |  |
| Food Insecurity X |  |  |  |  |  |  |  |  |  |
|  | AOR | CI | Sig. | AOR | CI | Sig. | AOR | CI | Sig. |
| Cisgender Heterosexual Female |  |  |  |  |  |  | 0.96 | (0.74,1.25) | 0.768 |
| LGBTQIA2+ |  |  |  |  |  |  | 0.59 | (0.42,0.82) | 0.001 |
| Race/Ethnicity: Asian |  |  |  |  |  |  | 0.69 | (0.22,2.17) | 0.517 |
| Race/Ethnicity: Black |  |  |  |  |  |  | 1.02 | (0.78,1.35) | 0.878 |
| Race/Ethnicity: Hispanic |  |  |  |  |  |  | 1.32 | (0.94,1.84) | 0.105 |
| Race/Ethnicity: Multiple |  |  |  |  |  |  | 1.02 | (0.66,1.58) | 0.942 |
| Race/Ethnicity: Other |  |  |  |  |  |  | 1.73 | (0.89,3.37) | 0.103 |
| Discrimination X |  |  |  |  |  |  |  |  |  |
| Cisgender Heterosexual Female |  |  |  |  |  |  | 0.96 | (0.69,1.32) | 0.778 |
| LGBTQIA2+ |  |  |  |  |  |  | 0.98 | (0.64,1.5) | 0.925 |
| Race/Ethnicity: Asian |  |  |  |  |  |  | 2.58 | (0.76,8.77) | 0.125 |
| Race/Ethnicity: Black |  |  |  |  |  |  | 1.47 | (0.99,2.17) | 0.053 |
| Race/Ethnicity: Hispanic |  |  |  |  |  |  | 0.82 | (0.48,1.38) | 0.445 |
| Race/Ethnicity: Multiple |  |  |  |  |  |  | 1.21 | (0.65,2.24) | 0.542 |
| Race/Ethnicity: Other |  |  |  |  |  |  | 0.65 | (0.24,1.78) | 0.397 |
| Neighborhood Social Cohesion X |  |  |  |  |  |  |  |  |  |
| Cisgender Heterosexual Female |  |  |  |  |  |  | 1.11 | (0.84,1.48) | 0.446 |
| LGBTQIA2+ |  |  |  |  |  |  | 1.41 | (0.94,2.10) | 0.090 |
| Race/Ethnicity: Asian |  |  |  |  |  |  | 3.95 | (1.29,12.11) | 0.015 |
| Race/Ethnicity: Black |  |  |  |  |  |  | 0.77 | (0.52,1.15) | 0.197 |
| Race/Ethnicity: Hispanic |  |  |  |  |  |  | 0.76 | (0.47,1.21) | 0.238 |
| Race/Ethnicity: Multiple |  |  |  |  |  |  | 0.97 | (0.53,1.79) | 0.922 |
| Race/Ethnicity: Other |  |  |  |  |  |  | 1.12 | (0.44,2.87) | 0.808 |
| Loneliness X |  |  |  |  |  |  |  |  |  |
| Cisgender Heterosexual Female |  |  |  |  |  |  | 0.44 | (0.32,0.61) | < 0.001 |
| LGBTQIA2+ |  |  |  |  |  |  | 0.22 | (0.14,0.35) | < 0.001 |
| Race/Ethnicity: Asian |  |  |  |  |  |  | 1.54 | (0.46,5.11) | 0.477 |
| Race/Ethnicity: Black |  |  |  |  |  |  | 0.67 | (0.42,1.07) | 0.094 |
|  | AOR | CI | Sig. | AOR | CI | Sig. | AOR | CI | Sig. |
| Race/Ethnicity: Hispanic |  |  |  |  |  |  | 1.25 | (0.71,2.22) | 0.437 |
| Race/Ethnicity: Multiple |  |  |  |  |  |  | 0.97 | (0.48,1.94) | 0.925 |
| Race/Ethnicity: Other |  |  |  |  |  |  | 1.19 | (0.40,3.54) | 0.751 |

### Social Position Main Effects

Compared to Cisgender Heterosexual Males, Cisgender Heterosexual Females and LGBTQIA2+ individuals are more than twice as likely to have an MDD diagnosis (AOR = 2.10 and 2.66, respectively, p < 0.001). Other social position factors also predict MDD. Low income (< $35,000) and less education (no college degree), respectively, also predict an MDD diagnosis compared to higher income and more education (AOR = 1.42 and 1.37, respectively, p < 0.001). Not owning a home and age above 65 years are similarly predictive of MDD, AOR = 1.13 and 1.11, respectively, p < 0.001. In our analysis, only Asian/Asian American race/ethnicity category differs significantly from White, with Asian/Asian American community members having an AOR = 0.10, p < 0.001. The AOR for MDD for all other race/ethnicity categories are statistically indistinguishable from that of Whites. In summary, relative to RQ1, our analysis shows that sexual and gender identity are strongly predictive of MDD, but that race and ethnicity are much less predictive of MDD.

### SDoH Main Effects

Higher Food Insecurity (AOR = 1.39, p = 0.009) and greater Loneliness (AOR = 15.09, p <0.001) both indicate a higher likelihood of MDD. Transience (AOR = 0.93, p < 0.001), Lack of Health Care Access (AOR = 1.35, p < 0.001), and having Health Insurance (AOR = 1.64, p < 0.001) also significantly predict MDD. Transience, with an AOR less than one, reduced the likelihood of having an MDD diagnosis; the other significant factors just mentioned increase the likelihood of an MDD diagnosis. In summary, relative to RQ2, our analysis indicates that Food Insecurity and Loneliness, as expected, are substantially connected to increased likelihood of MDD. Neither Discrimination nor Neighborhood Social Cohesion were significant predictors of MDD.

### Moderation Effects

Our analysis for RQ3 focuses on how race/ethnicity and gender and sexual identity interact with Food Insecurity, Discrimination, Neighbor Social Cohesion, and Loneliness to assess to what degree these four SDoH factor have differential impact on MDD for different identities. The interaction between Food Insecurity and Gender and Sexual Identity revealed a reduction in the likelihood of an MDD diagnosis for LGBTQIA2+ individuals (AOR = 0.59, p < 0.001). The interaction between Discrimination and Race/Ethnicity was not significant at 0.01, but it is worth noting that the AOR for Black community members was significant at 0.053 (with AOR = 1.47). The interaction between Neighborhood Social Cohesion and Race/Ethnic produced a significant result (AOR = 3.95, p < 0.001). Additionally, we also note that the interaction AOR for Neighborhood Social Cohesion and LGBTQIA2+ individuals was significant at 0.09 (AOR = 1.41). Last, the interaction between Loneliness and Race/Ethnicity revealed significant reductions in the likelihood of MDD in the interaction terms for both Cisgender Heterosexual Females and LGBTQIA2+ individuals, compared to Cisgender Heterosexual Males, with respective AOR values of 0.44 and 0.22, p < 0.001 for both. In summary, relative to RQ3, our analysis produced mixed support for differential SDoH effects on MDD for identities defined by race/ethnicity and gender/sexual orientation.

In the next section, we employ expected marginal means and interaction plots to support a deeper discussion of these analytical findings. This approach allows the effects of the various parameter estimates associated with our research questions to be highlighted by combining the main and interaction effects from stages 1, 2, and 3 while averaging across all other variables in the model.

## Discussion

Extant research has examined the roles of social position and social determinants of health on mental health outcomes. We add to this literature by focusing on major depressive disorder, investigating how race, ethnicity, gender, and sexual identity moderate the role of several social determinant domains on this common mental health condition. Our findings further illustrate the complexity and nuance associated with how the context of where and how people live their lives has significant differential impact on health outcomes.

Some of our results confirm long-standing relationships while elucidating detail about the effect on health. For example, controlling for social determinants of health and other social position factors, non-White community members have lower likelihood of MDD diagnosis. Asian community members are 62% less likely to be diagnosed with MDD (AOR = 0.38, p < 0.001), Black community members are 22% less likely to be diagnosed with MDD (AOR = 0.78, p < 0.001), Hispanic community members are 28% less likely to be diagnosed with MDD (AOR = 0.72, p < 0.001), and individuals who identify with multiple races/ethnicities are 27% less likely to be diagnosed with MDD (AOR = 0.73, p < 0.001), per Stage 2 regression results. However, in the case of Black individuals, the relationship between race and MDD diagnosis changes when moderation effects are included (AOR=1.00, p = 0.982), indicating the differential likelihood of MDD diagnosis between Black and White individuals is largely attributed to the unique moderation effects of social determinants of health on these two groups. It is important to reiterate that our analysis of MDD risk is based on a confirmed MDD diagnosis rather than self-report or perception of MDD. Because African Americans and Latinos, for example, are more likely to lack access to mental healthcare compared to Whites, the likelihood of receiving an MDD diagnosis may be lower. As a result, the prevalence of the disorder among some populations may be undercounted due to factors like these.

Figure 1 combines these effects visually to illustrate the differences between racial and ethnic groups as well as between gender and sexual identities. Differences in base rate, where perceived discrimination is low, are apparent. As perceived discrimination increases vastly different slopes are apparent until low sample power increases the variability to the extent that confidence intervals on the prediction overlap. It is worth noting that for the sake of parsimony, the plots in Figures 1 – 4 focus on the range 0 to 0.2 for the probability of an MDD diagnosis, causing the line for White community members to be truncated as the CI error bars expand for high level of loneliness.

**Figure 1.**
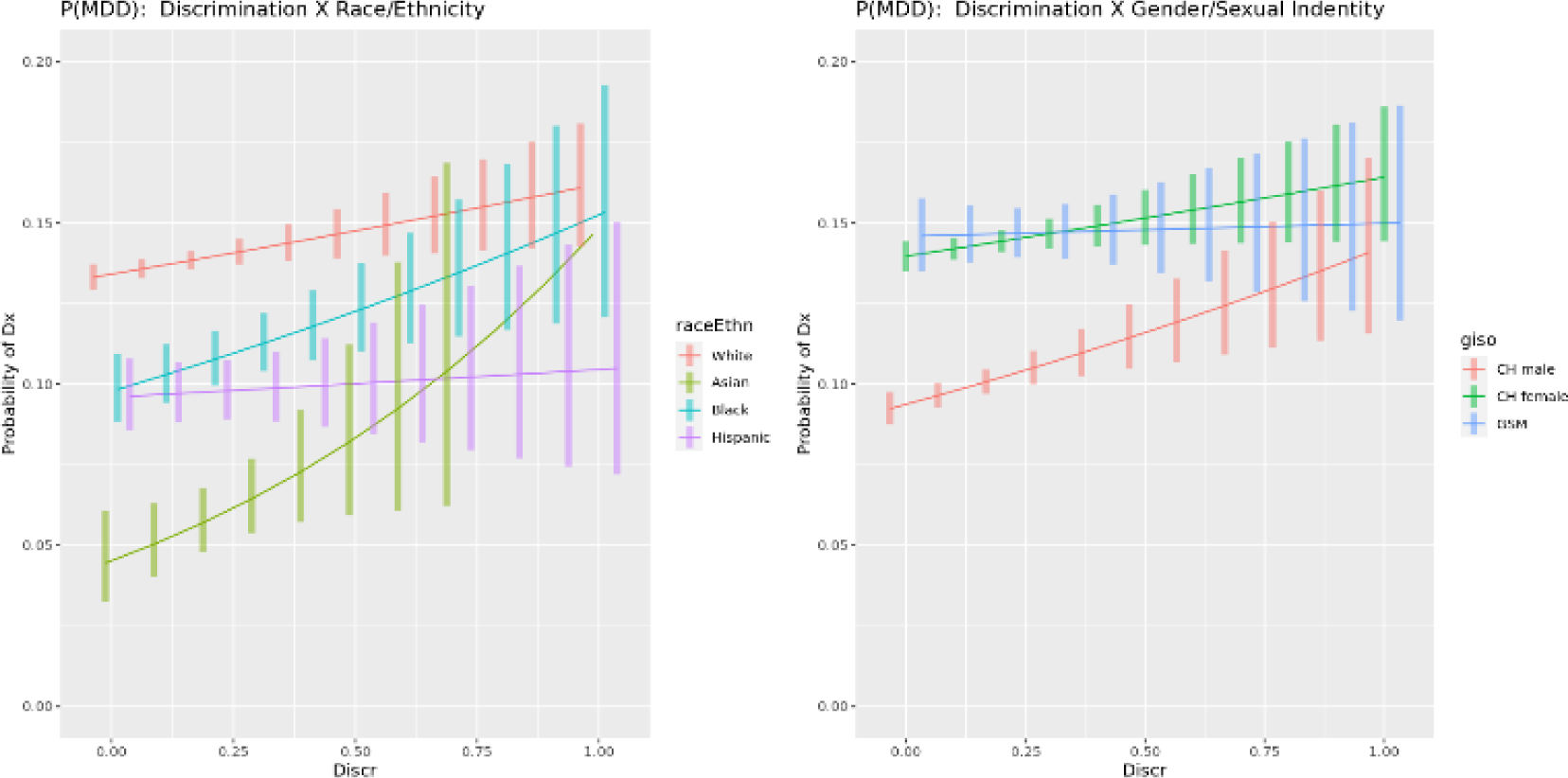
Interaction Plot for Discrimination. The interaction between the SDoH factor for discrimination, food insecurity, social cohesion, and loneliness versus the demographic factors for race and ethnicity and gender and sexual identity. The vertical axis is the predicted likelihood of an AoU participant in our analytical sample experiencing MDD. The horizontal axis for all SDoH factors has the range [0,1], where a higher value corresponds, respectively, with a participant’s neighborhood having more discrimination, greater food insecurity, greater social cohesion, and more loneliness.

**Figure 2.**
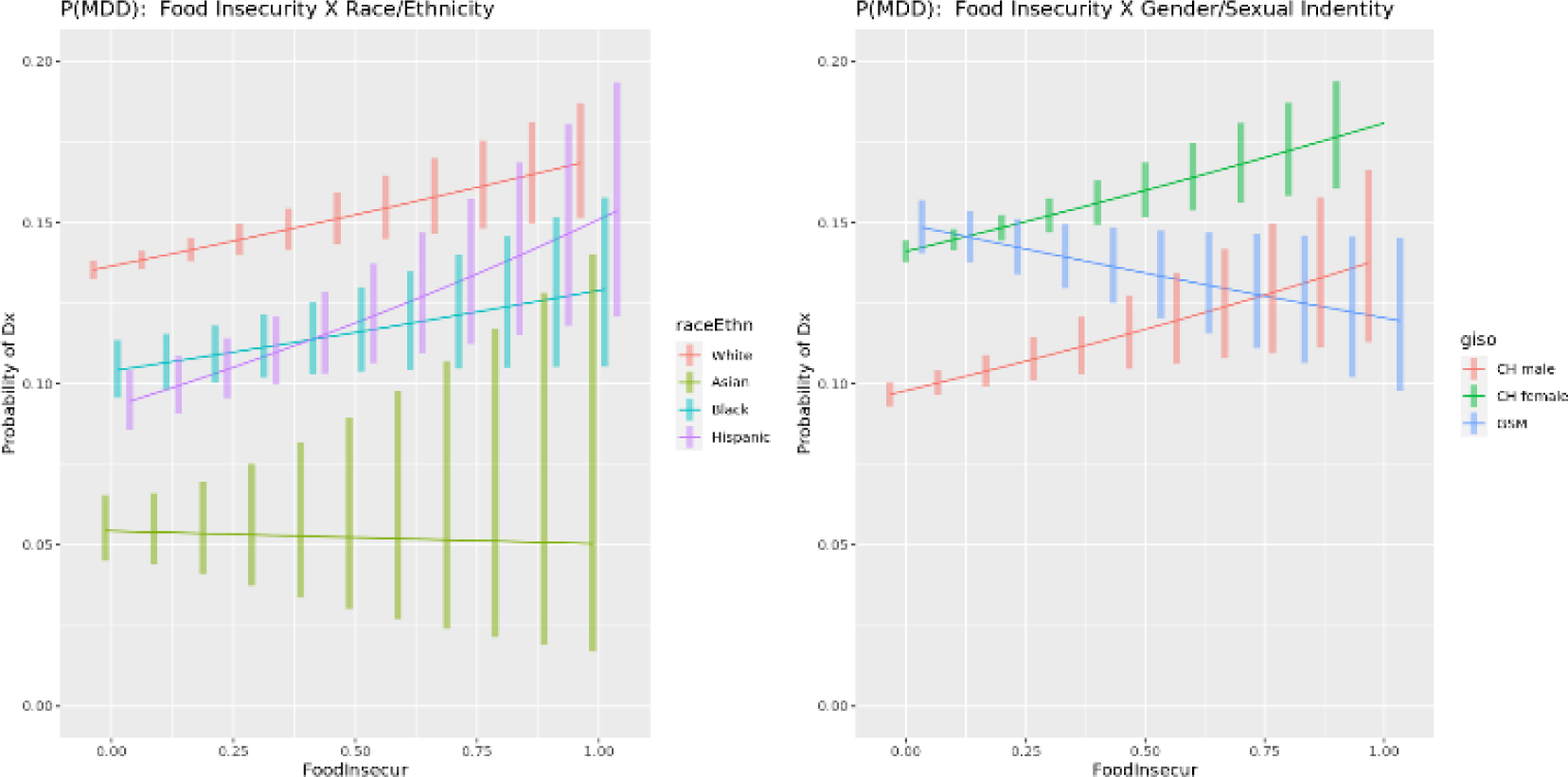
Interaction Plot for Food Insecurity. The interaction between the SDoH factor for discrimination, food insecurity, social cohesion, and loneliness versus the demographic factors for race and ethnicity and gender and sexual identity. The vertical axis is the predicted likelihood of an AoU participant in our analytical sample experiencing MDD. The horizontal axis for all SDoH factors has the range [0,1], where a higher value corresponds, respectively, with a participant’s neighborhood having more discrimination, greater food insecurity, greater social cohesion, and more loneliness.

**Figure 3.**
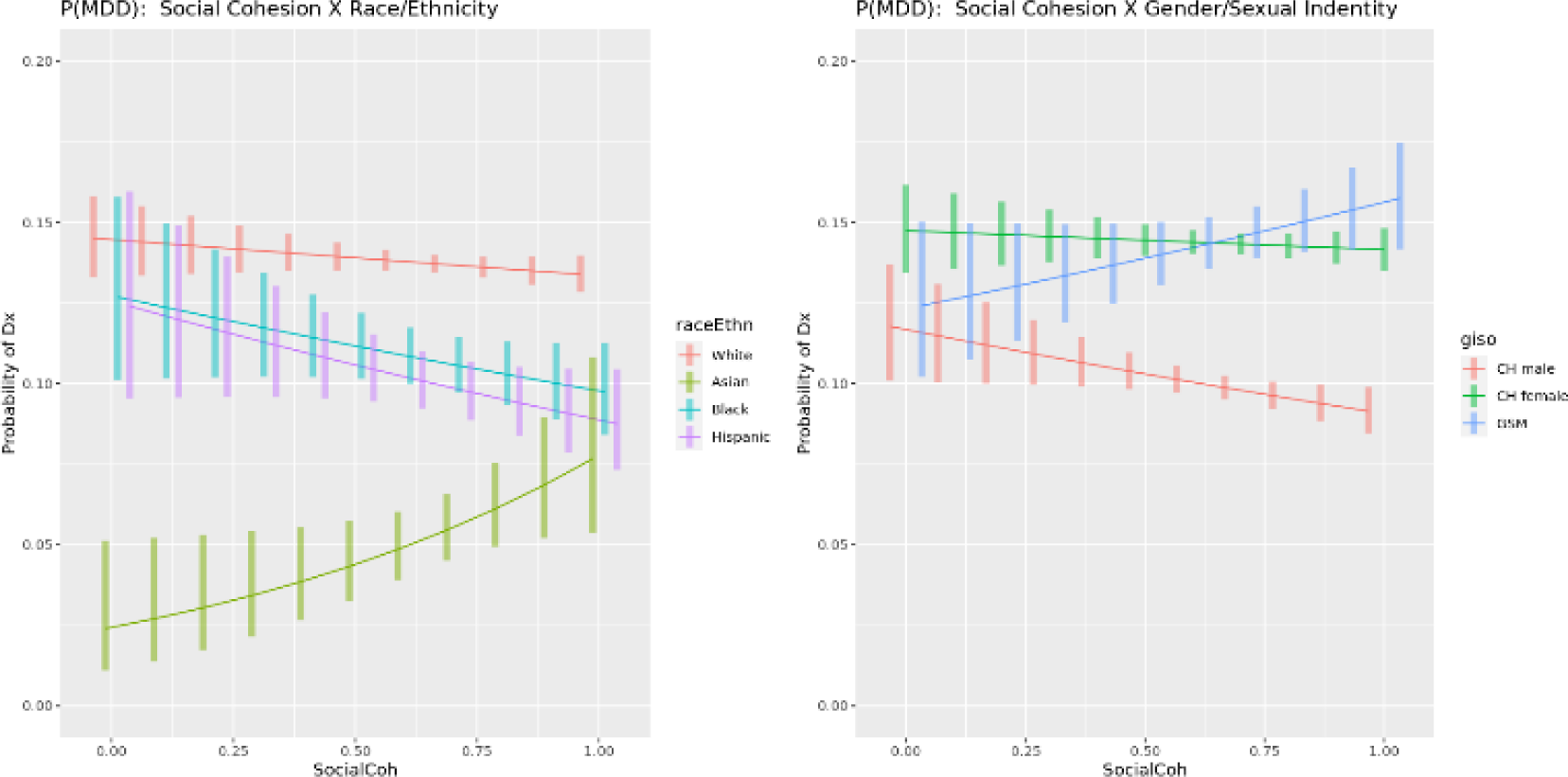
Interaction Plot for Social Cohesion. The interaction between the SDoH factor for discrimination, food insecurity, social cohesion, and loneliness versus the demographic factors for race and ethnicity and gender and sexual identity. The vertical axis is the predicted likelihood of an AoU participant in our analytical sample experiencing MDD. The horizontal axis for all SDoH factors has the range [0,1], where a higher value corresponds, respectively, with a participant’s neighborhood having more discrimination, greater food insecurity, greater social cohesion, and more loneliness.

**Figure 4.**
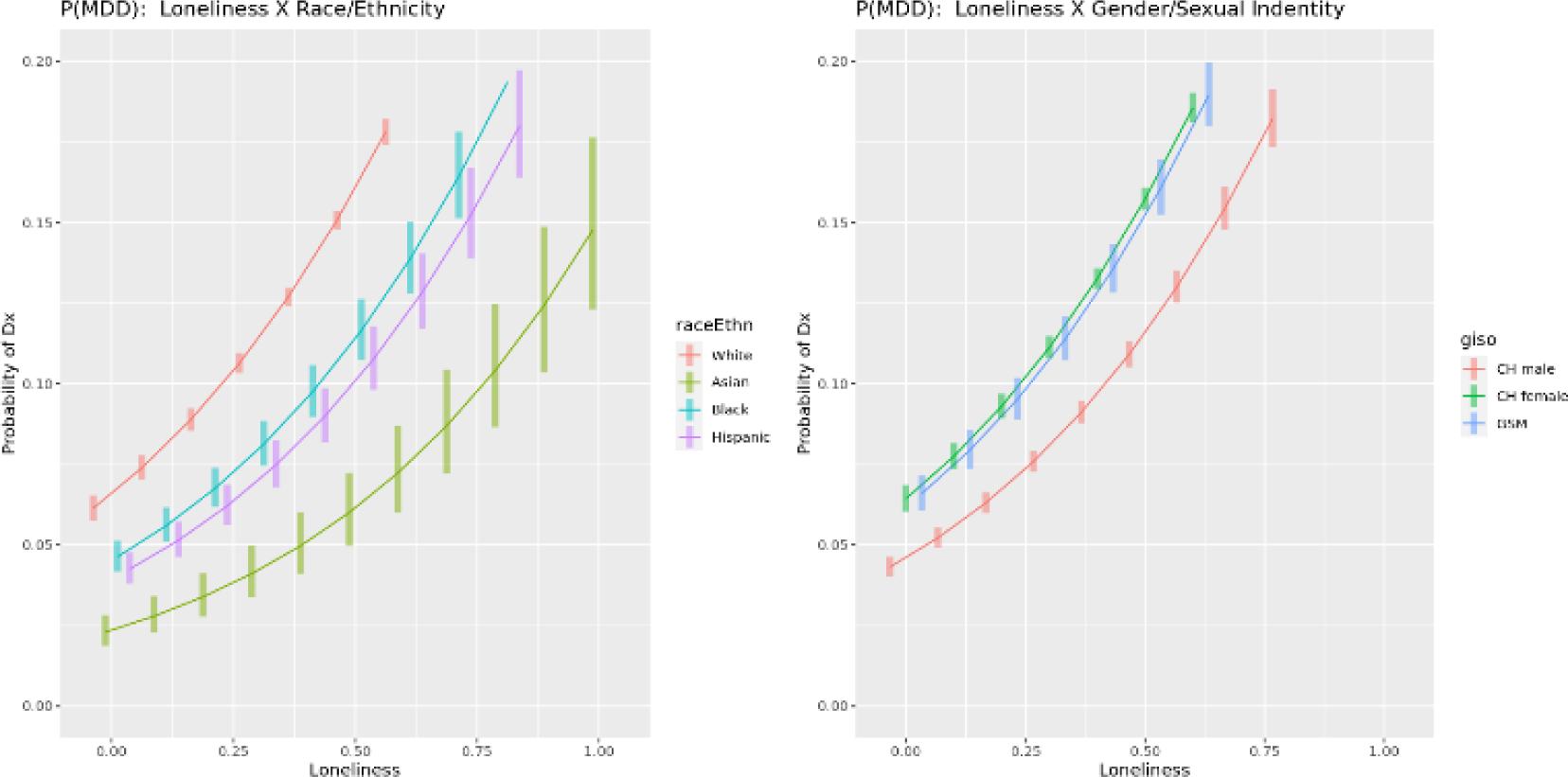
Interaction Plot for Loneliness. The interaction between the SDoH factor for discrimination, food insecurity, social cohesion, and loneliness versus the demographic factors for race and ethnicity and gender and sexual identity. The vertical axis is the predicted likelihood of an AoU participant in our analytical sample experiencing MDD. The horizontal axis for all SDoH factors has the range [0,1], where a higher value corresponds, respectively, with a participant’s neighborhood having more discrimination, greater food insecurity, greater social cohesion, and more loneliness.

As expected, loneliness is a substantial driver of depression, with increases to loneliness significantly increasing the likelihood of an MDD diagnosis as a main effect (AOR = 15.09, p < 0.001). However, our analysis indicates that loneliness affects cisgender heterosexual female community members and gender and sexually minoritized community members more weakly than cisgender heterosexual males (AOR = 0.44 and 0.22, p < 0.001, respectively). However, rather than indicate a protective effect of loneliness among these groups, the direction of these low odds ratios reflects two facts: (1) base rates of MDD are much higher in cishet females and LGTBQIA2+ individuals than cishet males and (2) base rates of loneliness are much higher in cishet females and LGTBQIA2+ individuals than cishet males. As a consequence, more of the effect of loneliness on MDD diagnosis is captured in the social position and loneliness main effects for cishet females and LGBTQIA2+ individuals compared to cishet males. In general, the relationship between loneliness and MDD diagnosis is very similar across racial/ethnic and gender/sexuality groups. Figure 4 illustrates the main effect of loneliness on depression at the same time that it elucidates the more modest moderation effects with race/ethnicity and gender/sexual identity.

Other results shed new light on less established moderation effects. For example, gender and sexually minoritized community members are much more likely to experience depression compared to cisgender heterosexual men (AOR = 2.66, p < 0.001). Drawing detail from Table 7, increasing neighborhood social cohesion does not alter the likelihood of depression, holding all other factors constant (AOR = 0.84, p = 0.181). But there is a weak moderation effect (AOR = 1.41, p = 0.090). As shown in Figure 3, while the base likelihood of an MDD diagnosis is undifferentiated at low levels of neighborhood social cohesion, distinctly different slopes are apparent as cohesion increases. In fact, LGBTQAI2+ community members stand out with a distinctly positive slope whereas cisgender heterosexual males and females have a flat or decreasing slope indicative of a protective factor for social cohesion relative to risk of depression. Similarly, Asian individuals stand out from White, Black, and Hispanic individuals with a distinctly positive moderation of social cohesion on likelihood of MDD diagnosis. Our analysis suggests that gender and sexual minorities and Asian community members may, relative to depression, be left out of the protective benefit of increased social cohesion.

Our results suggest the effect of discrimination on the likelihood of a diagnosis of major depressive disorder is different across racial/ethnic groups. The strongest signals we see for the effect of discrimination on MDD diagnosis are among Black and Asian groups. Our results suggest Black individuals reporting high discrimination are 47% more likely to be diagnosed with MDD than those reporting no discrimination (p = 0.053), and Asian individuals reporting high discrimination are 158% more likely to be diagnosed with MDD than those reporting no discrimination (p = 0.125). Visually, Figure 1 demonstrates the gradient of likelihood of MDD diagnosis with respect to discrimination is much larger in Black and Asian groups with respect to the reference level White group. The significance levels for these moderation effects are slightly above the conventional 0.05 cutoff, suggesting that further analysis with greater statistic power is needed to confirm our findings.

Increased levels of food insecurity differentially impact various community groups. As seen in Figure 2, generally, higher level of food insecurity corresponds with a higher likelihood of an MDD diagnosis, although slopes differ. Paradoxically, our analysis reveals a negative slope for LGBTQIA2+ community members, a finding that deserves additional research.

## Limitations

The All of Us dataset is a tremendous resource for the research community. Aptly named, it represents all of us, but it does so with social position representation that approximately mirrors that of the US population overall. As the current study examines, identity segments experience different health outcomes based on the context in which members of those identities live their lives. Some identities endure different effects from various SDoH, and the field would be better served by samples that provide sufficient power to make improved assessment of the moderation effects of SDoH on social position relative to health outcomes for marginalized community members. This is essential to improve understanding of disease prevalence and associated factors that will enable the delivery of more relevant and culturally competent mental health care.

The All of Us Research Program includes an expansive array of measurements providing support for extremely detailed analysis of health outcomes and the drivers of those outcomes. However, participants who enroll in All of Us typically complete The Basics, Lifestyle, and Overall Health Surveys upon enrollment. The Social Determinants of Health is administered as an optional follow-up survey to All of Us participants. Further, the Social Determinants of Health survey has only been available since November 2021, making it the newest follow-up survey offered to participants. Therefore, the Social Determinants of Health survey was not offered to most participants upon enrollment. Only 117,800 of the 413,460 individuals who have participated in the All of Us program have taken the Social Determinants of Health survey. Individuals who did not participate in the Social Determinants of Health survey were omitted from our analytical sample. Some of those who did participate in the Social Determinants of Health Survey, however, recorded null responses that disqualified them from inclusion in our analytical sample. Our final analytical sample includes 86,960 participants.

The All of Us dataset includes an expansive array of measurements that we have operationalized into a set of SDoH factors, described in detail above. We acknowledge that while we based our operationalization strategy based on a careful review of extant literature and SDoH screeners [18,34–37] and the sources of the instruments from which AoU drew the questions, other choices might have been made. See Bhavnani and colleagues (2023) for another approach to subtyping SDoH subdomains from the measurements provided by AoU [38].

The current study demonstrated the value of the AoU data in the study of how SDoH differentially drive health outcomes. It also provides a reminder that even larger datasets designed to represent the general population face substantial challenges for research focused on marginalized community segments and is a timely reminder that sampling plans are needed ensure sufficient statistical power to examine those most marginalized and underserved.

Despite these limitations, the All of Us program provides a research asset that is without peer. Future planning for the All of Us data might consider oversampling of participants within select identity segments in addition to expanding the size of the general population sample. Oversampling within the intersections of race/ethnicity and gender/sexual identity is an example. More specifically, increasing the sample of women and nonbinary community members of color would allow studies to elucidate ways to improve the delivery of care to community members who endure disproportionate impact from several SDoH factors on, in the current study, major depressive disorder.

## Conclusions

The current study employs the All of Us dataset to unpack the potentially intersecting effects of various SDoH factors to examine the different roles they play on depression based on race, ethnicity, gender, and sexual identity. We employ staged logistic regression examine main and moderating effects. Our analysis confirms the nuance and complexity of these relationships. We also use these analyses to outline future research to delve deeper into some of these findings. The current study demonstrated the value of the AoU data in the study of how various SDoH factors differentially drive health outcomes. It also provides a reminder that even larger datasets designed to represent the general population face substantial challenges for research focused on marginalized community segments and is a timely reminder that sampling plans are needed ensure sufficient statistical power to examine those most marginalized and underserved.

## Data Availability

The study uses the NIH All of Us Research Project as its source of data. These data and the computational notebooks we used are available through that public resource.

## Acknowledgement

The All of Us Research Program is supported by the National Institutes of Health, Office of the Director: Regional Medical Centers: 1 OT2 OD026549; 1 OT2 OD026554; 1 OT2 OD026557; 1 OT2 OD026556; 1 OT2 OD026550; 1 OT2 OD 026552; 1 OT2 OD026553; 1 OT2 OD026548; 1 OT2 OD026551; 1 OT2 OD026555; IAA #: AOD 16037; Federally Qualified Health Centers: HHSN 263201600085U; Data and Research Center: 5 U2C OD023196; Biobank: 1 U24 OD023121; The Participant Center: U24 OD023176; Participant Technology Systems Center: 1 U24 OD023163; Communications and Engagement: 3 OT2 OD023205; 3 OT2 OD023206; and Community Partners: 1 OT2 OD025277; 3 OT2 OD025315; 1 OT2 OD025337; 1 OT2 OD025276. In addition, the All of Us Research Program would not be possible without the partnership of its participants.

## Supplementary Materials

**Supplement 1:**
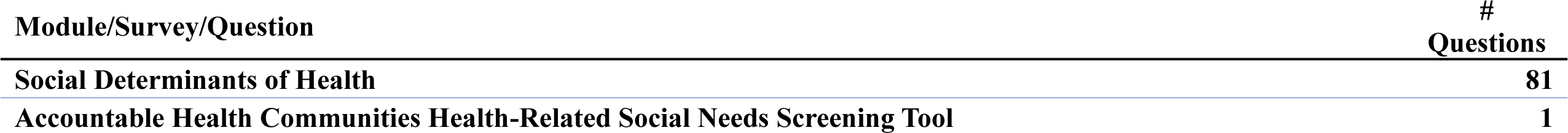
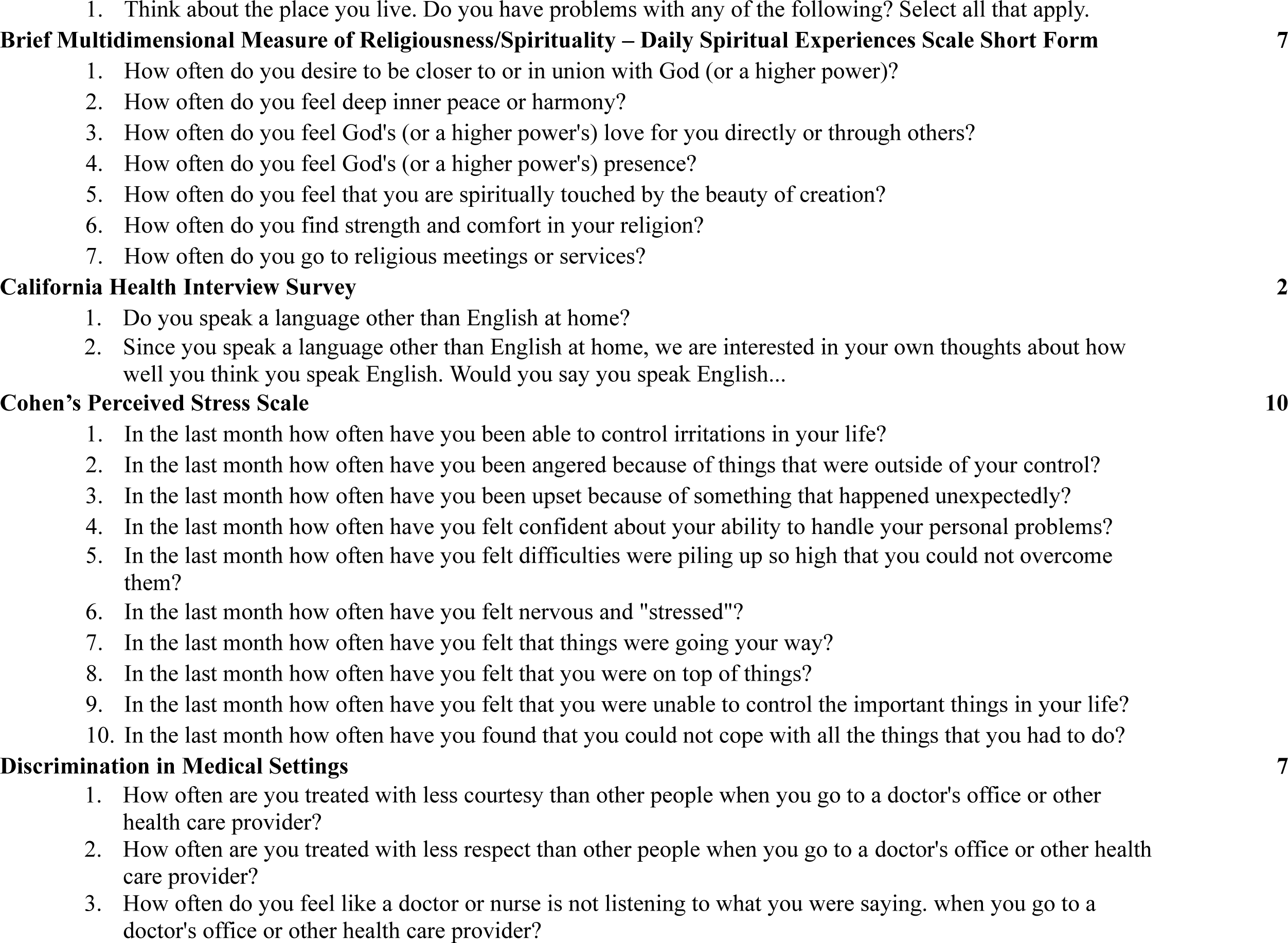
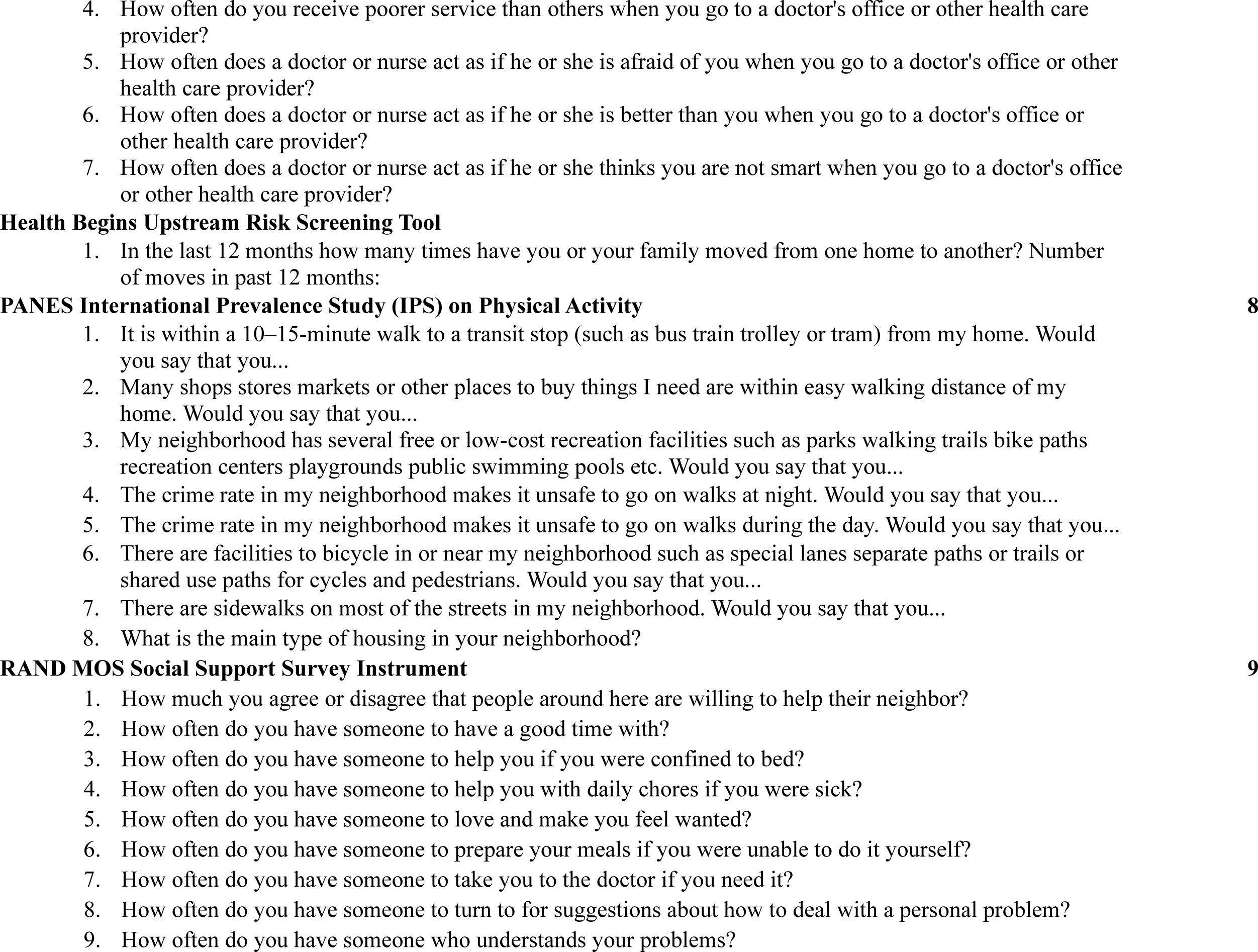
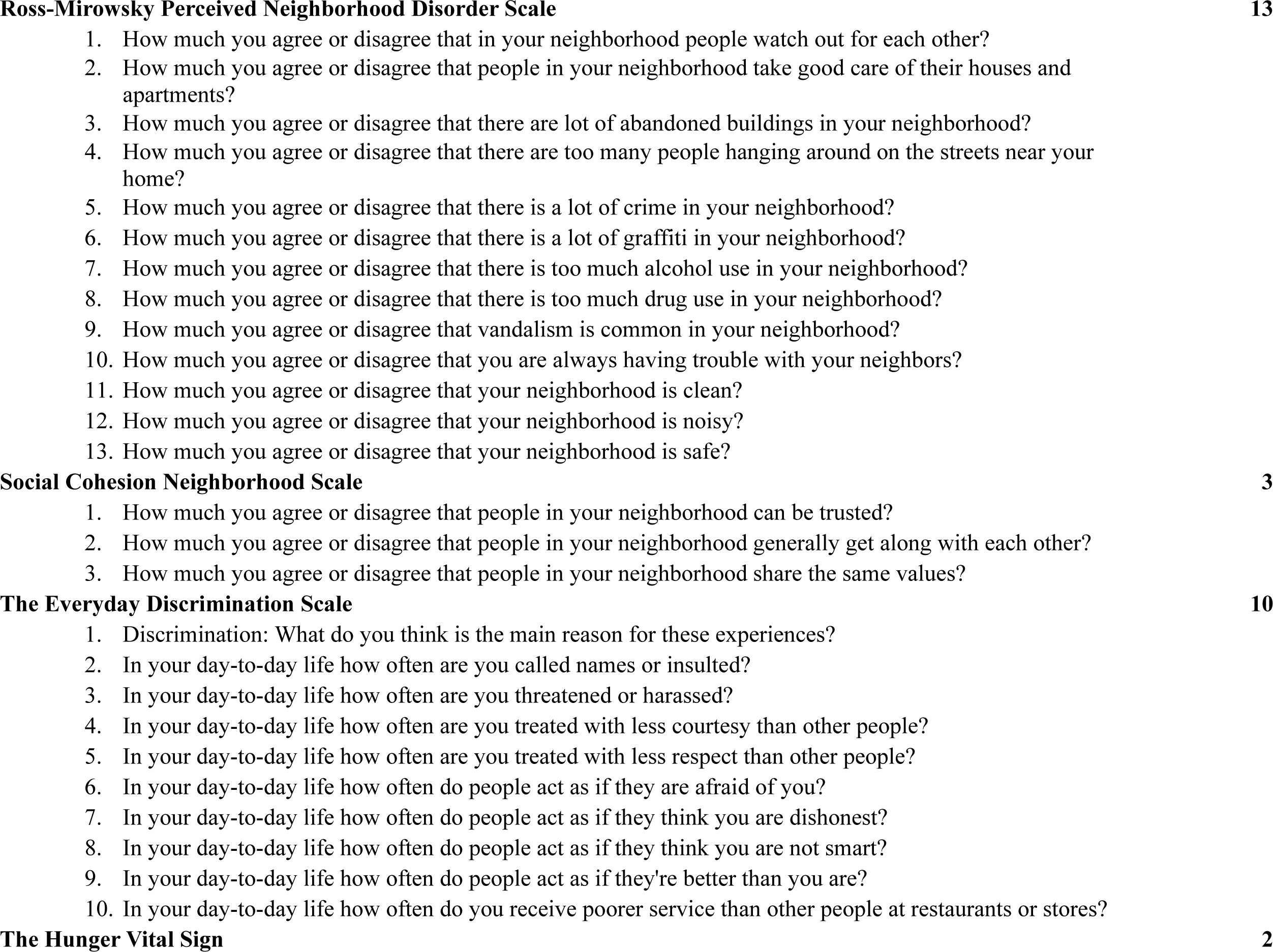
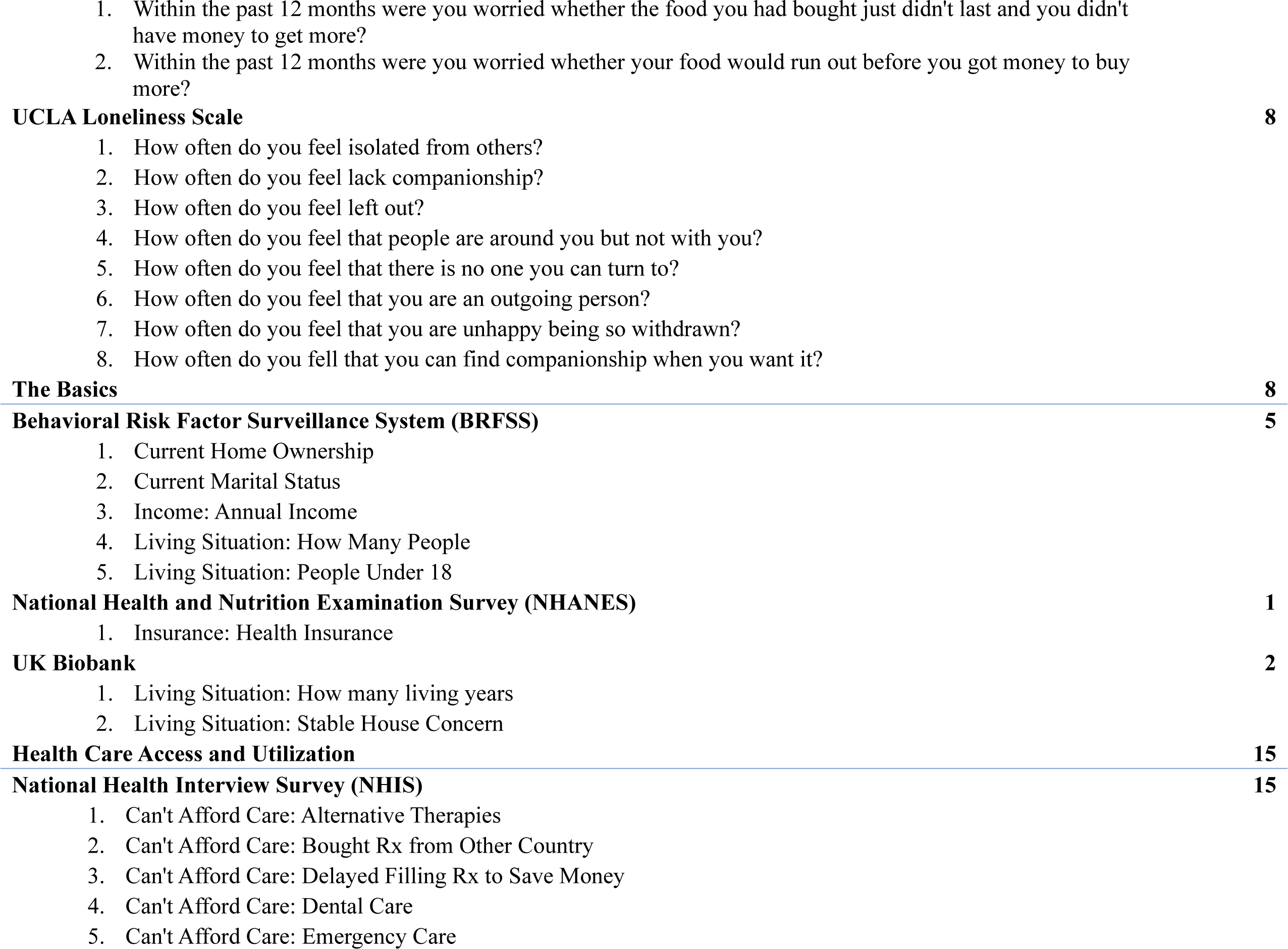
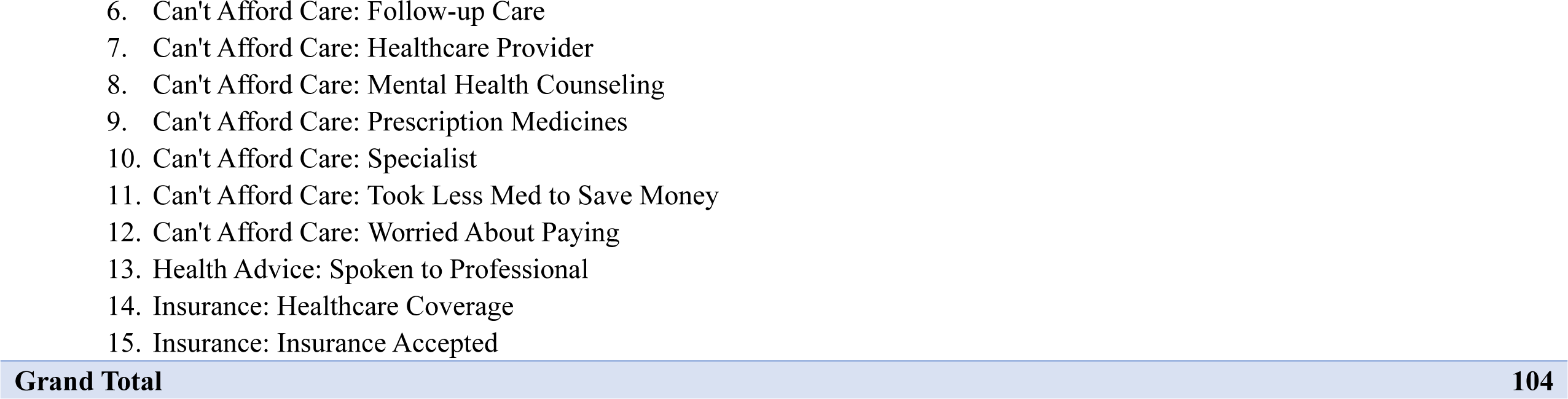
All of Us Measurements.

The All of Us SDoH survey draws from several instruments.

- Accountable Health Communities Health-Related Social Needs Screening Tool
- Behavioral Risk Factor Surveillance System (BRFSS)
- Brief Multidimensional Measure of Religiousness/Spirituality – Daily Spiritual Experiences Scale Short Form
- California Health Interview Survey
- Cohen’s Perceived Stress Scale
- Discrimination in Medical Settings
- Health Begins Upstream Risk Screening Tool
- National Health and Nutrition Examination Survey (NHANES)
- National Health Interview Survey (NHIS)
- PANES International Prevalence Study (IPS) on Physical Activity
- RAND MOS Social Support Survey Instrument
- Ross-Mirowsky Perceived Neighborhood Disorder Scale
- Social Cohesion Neighborhood Scale
- The Everyday Discrimination Scale
- The Hunger Vital Sign
- UCLA Loneliness Scale
- UK Biobank.

The All of Us Social Determinants of Health Survey questions were aggregated from these validated instruments designed to measure specific social determinants of health. The items “health care access and coverage” are not included in the All of Us Social Determinants of Health survey. These items are covered in The Basics and Health Care Access and Utilization surveys, which were administered before the Social Determinants of Health Survey. We combine selected health care questions from these other surveys with the Social Determinants of Health survey to form the corpus of questions for our analysis of social determinants of health. Listed below are the modules and the questions utilized in the current study.

**Supplement 2:**
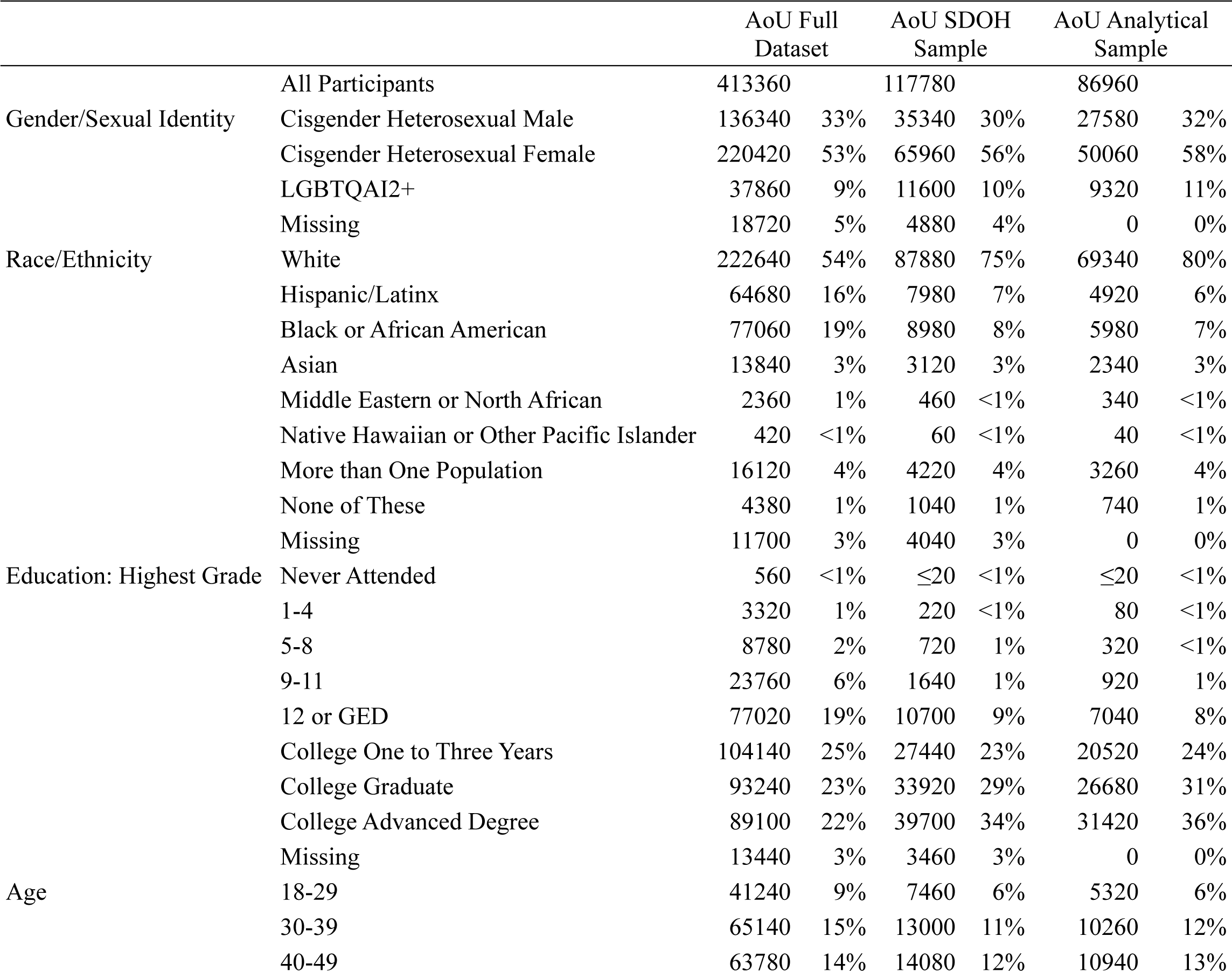
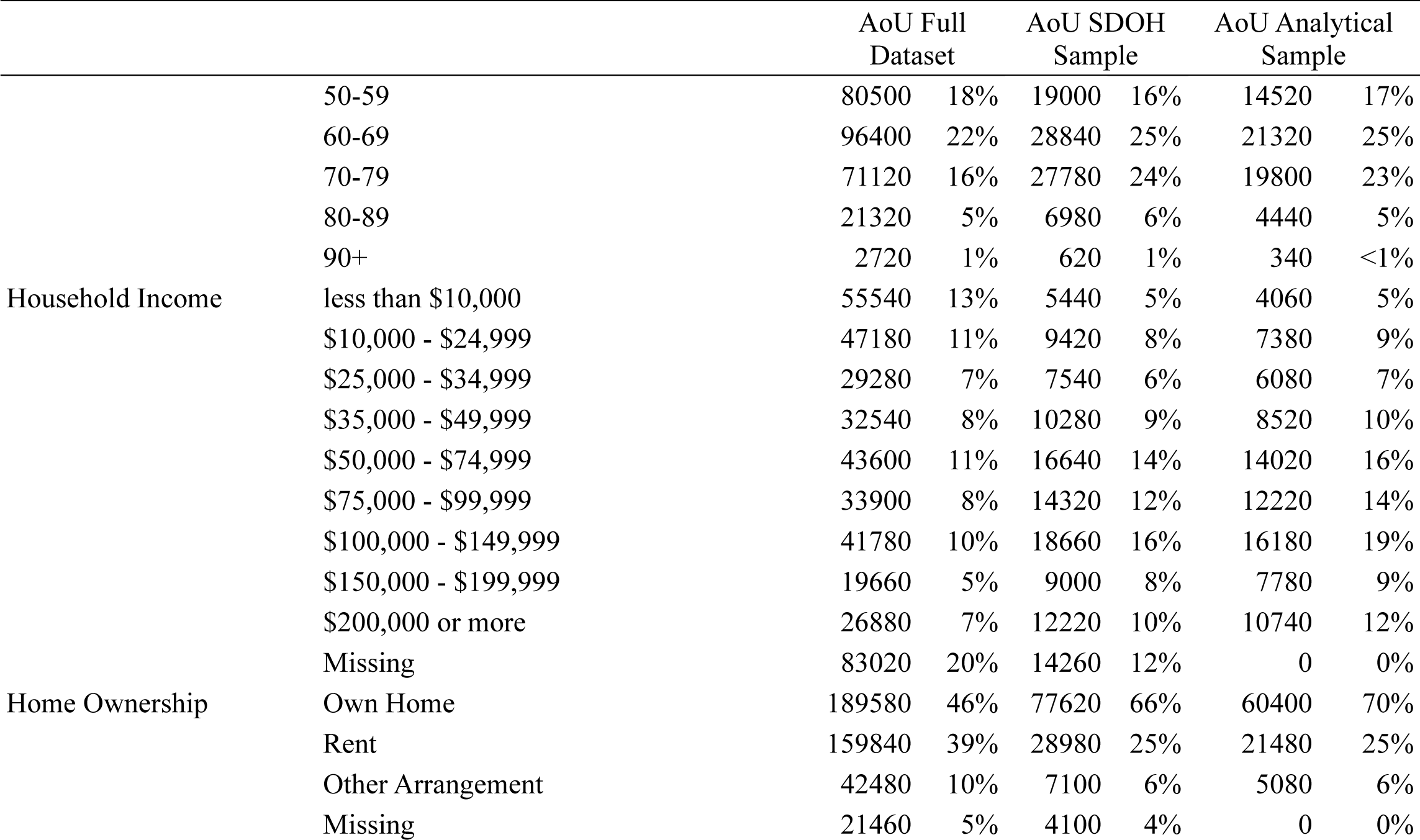
Social Position Distribution.

## Notes

### Competing Interest Statement

The authors have declared no competing interest.

### Funding Statement

This research was supported by institutional funding at The University of Texas at Austin under the Good Systems project AI and the Future of Racial Justice and the IC2 Institute's project Delivering Equitable Healthcare.

## References

1. Commission on Social Determinants of Health. Closing the gap in a generation : health equity through action on the social determinants of health : final report of the commission on social determinants of health. Combler Fossé En Une Génér Instaur Léquité En Santé En Agissant Sur Déterm Sociaux Santé Rapp Final Comm Déterm Sociaux Santé. 2008; 247. Available: https://apps.who.int/iris/handle/10665/43943

2. Social determinants of health. [cited 1 Nov 2023]. Available: https://www.who.int/health-topics/social-determinants-of-health

3. Social Determinants of Health - Healthy People 2030 | health.gov. [cited 1 Nov 2023]. Available: https://health.gov/healthypeople/priority-areas/social-determinants-health

4. Hill-Briggs F, Adler NE, Berkowitz SA, Chin MH, Gary-Webb TL, Navas-Acien A, et al. Social Determinants of Health and Diabetes: A Scientific Review. Diabetes Care. 2021;44: 258–279. doi:10.2337/dci20-0053

5. Pikhart H, Pikhartova J. The Relationship Between Psychosocial Risk Factors and Health Outcomes of Chronic Diseases: A Review of the Evidence for Cancer and Cardiovascular Diseases. Copenhagen: WHO Regional Office for Europe; 2015. Available: http://www.ncbi.nlm.nih.gov/books/NBK379457/

6. Silva M, Loureiro A, Cardoso G. Social determinants of mental health: a review of the evidence. Eur J Psychiatry. 2016;30: 259–292. Available: https://scielo.isciii.es/scielo.php?script=sci_abstract&pid=S0213-61632016000400004&lng=es&nrm=iso&tlng=en

7. Mental Health - Household Pulse Survey - COVID-19. 11 Oct 2023 [cited 31 Oct 2023]. Available: https://www.cdc.gov/nchs/covid19/pulse/mental-health.htm

8. What Makes All of Us Different. In: All of Us Research Program | NIH [Internet]. 15 Sep 2020 [cited 1 Nov 2023]. Available: https://allofus.nih.gov/about/program-overview/what-makes-all-us-different

9. Penninx BWJH, Benros ME, Klein RS, Vinkers CH. How COVID-19 shaped mental health: from infection to pandemic effects. Nat Med. 2022;28: 2027–2037. doi:10.1038/s41591-022-02028-2

10. Gong Y, Liu X, Zheng Y, Mei H, Que J, Yuan K, et al. COVID-19 Induced Economic Slowdown and Mental Health Issues. Front Psychol. 2022;13. Available: https://www.frontiersin.org/articles/10.3389/fpsyg.2022.777350

11. Bucciarelli V, Nasi M, Bianco F, Seferovic J, Ivkovic V, Gallina S, et al. Depression pandemic and cardiovascular risk in the COVID-19 era and long COVID syndrome: Gender makes a difference. Trends Cardiovasc Med. 2022;32: 12–17. doi:10.1016/j.tcm.2021.09.009

12. Bourmistrova NW, Solomon T, Braude P, Strawbridge R, Carter B. Long-term effects of COVID-19 on mental health: A systematic review. J Affect Disord. 2022;299: 118–125. doi:10.1016/j.jad.2021.11.031

13. Mezzina R, Gopikumar V, Jenkins J, Saraceno B, Sashidharan SP. Social Vulnerability and Mental Health Inequalities in the “Syndemic”: Call for Action. Front Psychiatry. 2022;13. Available: https://www.frontiersin.org/articles/10.3389/fpsyt.2022.894370

14. Walker ER, McGee RE, Druss BG. Mortality in Mental Disorders and Global Disease Burden Implications: A Systematic Review and Meta-analysis. JAMA Psychiatry. 2015;72: 334. doi:10.1001/jamapsychiatry.2014.2502

15. Powell-Wiley TM, Baumer Y, Baah FO, Baez AS, Farmer N, Mahlobo CT, et al. Social Determinants of Cardiovascular Disease. Circ Res. 2022;130: 782–799. doi:10.1161/CIRCRESAHA.121.319811

16. Broadhead WE. Depression, Disability Days, and Days Lost From Work in a Prospective Epidemiologic Survey. JAMA J Am Med Assoc. 1990;264: 2524. doi:10.1001/jama.1990.03450190056028

17. Briley M, Lépine. The increasing burden of depression. Neuropsychiatr Dis Treat. 2011; 3. doi:10.2147/NDT.S19617

18. Alegría M, NeMoyer A, Falgàs Bagué I, Wang Y, Alvarez K. Social Determinants of Mental Health: Where We Are and Where We Need to Go. Curr Psychiatry Rep. 2018;20: 95. doi:10.1007/s11920-018-0969-9

19. Gnanapragasam SN, Astill Wright L, Pemberton M, Bhugra D. Outside/inside: social determinants of mental health. Ir J Psychol Med. 2021; 1–11. doi:10.1017/ipm.2021.49

20. Compton MT, Shim RS. The Social Determinants of Mental Health. FOCUS. 2015;13: 419–425. doi:10.1176/appi.focus.20150017

21. Jeste DV, Pender VB. Social Determinants of Mental Health: Recommendations for Research, Training, Practice, and Policy. JAMA Psychiatry. 2022;79: 283–284. doi:10.1001/jamapsychiatry.2021.4385

22. Fisher M, Baum F. The Social Determinants of Mental Health: Implications for Research and Health Promotion. Aust N Z J Psychiatry. 2010;44: 1057–1063. doi:10.3109/00048674.2010.509311

23. Reeder N, Tapanee P, Persell A, Tolar-Peterson T. Food Insecurity, Depression, and Race: Correlations Observed Among College Students at a University in the Southeastern United States. Int J Environ Res Public Health. 2020;17: 8268. doi:10.3390/ijerph17218268

24. Miller GH, Marquez-Velarde G, Lindstrom E-D, Keith VM, Brown LE. Neighborhood cohesion and psychological distress across race and sexual orientation. SSM - Popul Health. 2022;18: 101134. doi:10.1016/j.ssmph.2022.101134

25. Swann G, Stephens J, Newcomb ME, Whitton SW. Effects of sexual/gender minority- and race-based enacted stigma on mental health and substance use in female assigned at birth sexual minority youth. Cultur Divers Ethnic Minor Psychol. 2020;26: 239–249. doi:10.1037/cdp0000292

26. Khan M, Ilcisin M, Saxton K. Multifactorial discrimination as a fundamental cause of mental health inequities. Int J Equity Health. 2017;16: 43. doi:10.1186/s12939-017-0532-z

27. Adler NE, Epel ES, Castellazzo G, Ickovics JR. Relationship of subjective and objective social status with psychological and physiological functioning: Preliminary data in healthy, White women. Health Psychol. 2000;19: 586–592. doi:10.1037/0278-6133.19.6.586

28. Lindemann K. THE IMPACT OF OBJECTIVE CHARACTERISTICS ON SUBJECTIVE SOCIAL POSITION. TRAMES. 2007;XI: 54–68. Available: https://www.ceeol.com/search/article-detail?id=164149

29. Avlund K, Holstein BE, Osler M, Damsgaard MT, Holm-Pedersen P, Rasmussen NK. Social position and health in old age: the relevance of different indicators of social position. Scand J Public Health. 2003;31: 126–136. doi:10.1080/14034940210134130

30. The All of Us Research Program Investigators. The “All of Us” Research Program. N Engl J Med. 2019;381: 668–676. doi:10.1056/NEJMsr1809937

31. Mapes BM, Foster CS, Kusnoor SV, Epelbaum MI, AuYoung M, Jenkins G, et al. Diversity and inclusion for the All of Us research program: A scoping review. PLOS ONE. 2020;15: e0234962. doi:10.1371/journal.pone.0234962

32. Cronin RM, Jerome RN, Mapes B, Andrade R, Johnston R, Ayala J, et al. Development of the Initial Surveys for the All of Us Research Program. Epidemiol Camb Mass. 2019;30: 597–608. doi:10.1097/EDE.0000000000001028

33. Barr PB, Bigdeli TB, Meyers JL. Prevalence, Comorbidity, and Sociodemographic Correlates of Psychiatric Diagnoses Reported in the All of Us Research Program. JAMA Psychiatry. 2022;79: 622– 628. doi:10.1001/jamapsychiatry.2022.0685

34. Moen M, Storr C, German D, Friedmann E, Johantgen M. A Review of Tools to Screen for Social Determinants of Health in the United States: A Practice Brief. Popul Health Manag. 2020;23: 422–429. doi:10.1089/pop.2019.0158

35. Berkowitz RL, Bui L, Shen Z, Pressman A, Moreno M, Brown S, et al. Evaluation of a social determinants of health screening questionnaire and workflow pilot within an adult ambulatory clinic. BMC Fam Pract. 2021;22: 256. doi:10.1186/S12875-021-01598-3

36. O’Gurek DT, Henke C. A practical approach to screening for social determinants of health. Fam Pract Manag. 2018;25: 7–11. Available: https://www.aafp.org/pubs/fpm/issues/2018/0500/p7.html

37. Friedman NL, Banegas MP. Toward Addressing Social Determinants of Health: A Health Care System Strategy. Perm J. 2018;22: 18–095. doi:10.7812/TPP/18-095

38. Bhavnani SK, Zhang W, Bao D, Raji M, Ajewole V, Hunter R, et al. Subtyping Social Determinants of Health in All of Us: Opportunities and Challenges in Integrating Multiple Datatypes for Precision Medicine. medRxiv; 2023. p. 2023.01.27.23285125. doi:10.1101/2023.01.27.23285125

39. Dunlop DD, Song J, Lyons JS, Manheim LM, Chang RW. Racial/Ethnic Differences in Rates of Depression Among Preretirement Adults. Am J Public Health. 2003;93: 1945–1952. doi:10.2105/AJPH.93.11.1945

40. Jeste DV, Pender VB. Social Determinants of Mental Health: Recommendations for Research, Training, Practice, and Policy. JAMA Psychiatry. 2022;79: 283. doi:10.1001/jamapsychiatry.2021.4385

